# Adolescent Cardiorespiratory Fitness and Future Risk of Depression and Antidepressants: A Nationwide Cohort Study with Sibling Comparators

**DOI:** 10.1101/2025.02.07.25321835

**Authors:** Marcel Ballin, Örjan Ekblom, Anna Nordström, Viktor H. Ahlqvist, Peter Nordström

## Abstract

**Objective:** To examine the association between adolescent cardiorespiratory fitness and future risk of incident depression and dispensation of antidepressants, while addressing the role of familial confounding.

**Methods:** A nationwide cohort study with registry-linkage and incorporating full sibling comparators, based on Swedish men who participated in mandatory military conscription examinations between 1972 and 1995. Cardiorespiratory fitness was estimated using a maximal ergometer bicycle test. Depression diagnosis in inpatient or specialised outpatient care, and dispensation of antidepressants until 31 December 2023, was ascertained using nationwide registers.

**Results:** A total of 1 013 885 men (mean age 18.3 years), of which 410 198 were full siblings, were followed until a median age of 56.8 years, during which 47 283 were diagnosed with depression and 237 409 were dispensed antidepressants at least once (18 150 and 93 150 among siblings). In cohort analysis, after controlling for observed confounders, the highest decile of fitness had lower risks of depression (hazard ratio [HR] 0.54; 95% CI 0.52, 0.57) and antidepressants (HR 0.63; 0.62, 0.65) compared to the lowest decile, with a difference in the standardised cumulative incidence at age 65 of −3.9% and −12.3% respectively. In sibling-comparison analyses accounting for unobserved familial confounders, the associations attenuated for both depression (HR 0.67, 0.59 to 0.75; incidence difference −2.4%) and antidepressants (HR 0.76, 0.72 to 0.80; incidence difference −7.2%). Hypothetically shifting everyone to the highest decile of fitness was associated with a preventable fraction of 29.1% for depression and 17.8% for antidepressants in cohort analysis, which attenuated to 17.6% and 10.4% in sibling-comparisons.

**Conclusions:** Higher levels of adolescent cardiorespiratory fitness were associated with a lower risk of future depression diagnosis and dispensation of antidepressants, but the associations may be overstated due to familial confounding. These findings highlight the importance of triangulation to better understand the link between adolescent cardiorespiratory fitness and future risk of depressive disorders.

**Key points:**

- In this nationwide cohort study of more than 1 million men, of which more than 400,000 were full siblings, higher levels of adolescent cardiorespiratory fitness were associated with lower risks of future depression and dispensation of antidepressants even up to five decades later.
- However, after controlling for unobserved confounders shared between full siblings (i.e., behavioural, environmental, and genetic factors), the magnitude of the associations attenuated by up to 40%, suggesting that the benefits associated with fitness may be overstated due to familial confounding.
- Monitoring cardiorespiratory fitness among adolescents could offer insights into their future risk of depression, but the causal relationship may be overestimated in traditional studies, underscoring the need for further causal analyses to triangulate results and yield more robust estimates.

## 1. Introduction

The prevalence of depression and other mental health problems remains at a staggering level, presenting a significant global health burden.[1, 2] While modifiable factors such as physical exercise can play a role in treatment outcomes when combined with standard care,[3] attention has also turned to whether exercise could prevent the onset of mental health problems, including depression.[4]

Recent studies have found a protective association between cardiorespiratory fitness – a marker of long-term physical exercise and genetic predisposition[5, 6] - and incident depression and use of antidepressants,[7–13] with proposed mechanisms including reduced stress,[14] enhanced psychosocial risk factors,[15] and effects on neurotrophic processes, monamine metabolism, and inflammation.[16] However, whether these epidemiological associations reflect causal relationships (i.e., increasing fitness will lower the risk of depression) or whether fitness is a marker of a broader, well-balanced lifestyle that coincides with better mental health, remains unclear. This distinction has important implications, where if causal, the findings could inform targeted interventions to reduce the incidence of depression. Alternatively, if primarily servering as a marker, the emphasis may need to shift toward assessing fitness in screening purposes. Unfortunately, disentangling a potential causal role of cardiorespiratory fitness in depression etiology has proven challenging because the field relies heavily on observational evidence – much of which lacks an adequate control for key biases, including unobserved familial confounding factors. Moreover, the abundance of studies have focused on middle-aged individuals, but very few have investigated the role of cardiorespiratory fitness in adolescence for future risk of depression;[9] which could have public health relevance as fitness levels track from adolescence into adulthood.[17]

In this study, we leverage Swedish conscription data on standardised cardiorespiratory fitness assessments of nearly the entire male adolescent population, linked to nationwide healthcare and population registries, to investigate the association between adolescent cardiorespiratory fitness and future risk of incident depression and antidepressant dispensations in over 1 million young men. To distinguish between a screening effect and a putative causal relationship, we compare the standard cohort analysis with estimates derived using sibling-comparison analysis involving over 400,000 full siblings, thus accounting for all shared behavioural, environmental, and genetic factors.[18]

## 2. Methods

### 2.1 Study design

We designed a cohort study by cross-linking data from Swedish registries using the Personal Identification Number, which is unique for all Swedish residents.[19] The study was approved by the Regional Ethical Review Board in Umeå and later by the Swedish Ethical Review Authority (no. 2010-113-31M), who waived the need for informed consent.[20] The study is reported according to the RECORD guidelines.[21]

### 2.2 Databases

The eligible study population was obtained from the Swedish Military Service Conscription Register,[22] and was based on all men who participated in military conscription examinations between 1972 and 1995. During this period, conscription around the age of 18 years was mandatory for all Swedish men, with few exemptions (approximately 90% population coverage).[22] Full siblings were identified using the Multi-Generation Register, ensuring accurate linkage of familial relationships.[23] Using the National Patient Register,[24] we collected data on depression diagnoses from inpatient and specialist outpatient care. Using the Prescribed Drug register, we collected data on antidepressive medications dispensed at pharmacies.[25] Deaths were tracked using the Cause of Death Register,[23] and emigration and socioeconomic data were obtained from Statistics Sweden.[23] Overall, these data cover the entire Swedish population and are mandated by law.

### 2.3 Derivation of study population

A total of 1 249 131 men were conscripted between 1972 and 1995. From these, we excluded 33 645 (2.7%) with missing fitness data, 182 856 (14.6%) with missing covariate data, and 18 745 (1.5%) with extreme values (detailed below). This resulted in 1 013 885 individuals being included in the cohort analysis (81.2% retained). Among these, 410 198 were full siblings from 188 946 families and were included in the sibling analysis (supplemental figure 1).

### 2.4 Estimation of cardiorespiratory fitness

Cardiorespiratory fitness was estimated at conscription using a maximal ergometer bicycle test according to a standardised procedure.[22] In short, conscripts performed the test following presentation of normal electrocardiography. The conscript began cycling for 5 minutes at 60 to 70 revolutions per minute at a low level of external resistance that was predetermined according to body weight. From thereon, resistance was gradually increased by 25 watts per minute until exhaustion. The results of the tests were recorded as Watt maximum (Wmax), which served as the exposure in the primary analysis. Wmax has been found to correlate strongly with measured maximal oxygen uptake (gold standard).[26] We excluded conscripts with extremely low values (<100 Wmax or ≥999 Wmax).

### 2.5 Ascertainment of outcomes

Depression diagnosis and dispensation of antidepressants were studied as two separate outcomes. Diagnosis of depression was defined as the date of first hospitalisation or visit in specialised outpatient care in the National Patient Register, recorded from 1997 until 31 December 2023 using the International Classification of Diseases 10^th^ revision code F32 in a primary or secondary position. The diagnostic validity of inpatient diagnoses in the National Patient Register is typically high (positive predictive values between 85% and 95% for most diagnoses), although sensitivity is typically lower.[24] The previous validation study did not describe data for depression, but a recent sub-study found fair to moderate agreement between depression diagnoses in the National Patient Register and another clinical register.[27] Dispensation of antidepressants was defined as the date of first dispensation, recorded from July 2005 until 31 December 2023 using the Anatomical Therapeutic Chemical code N06A in the Prescribed Drug Register. Dispensation of antidepressants would likely capture both some depression diagnosed in primary care (as primary care is not covered in the National Patient Register), as well as other milder psychiatric disorders such as anxiety disorders.

### 2.6 Covariates

From the Swedish Military Service Conscription Register, we collected data on age at conscription, year of conscription, objectively measured body mass index (BMI, kg/m^2^), and results from an IQ-test, as described previously.[22] Conscripts with extreme BMI values (≤15 and ≥60 kg/m^2^) were excluded. IQ was assessed using a standardised test battery involving subtests covering logical, verbal, and visospatial abilities, as well as a technical test. The individual subtests are summed to a total score, which is standardised by conscription year. We centered the distribution to a mean of 100 and a standard deviation of 15. From Statistics Sweden,[28] we obtained data on the socioeconomic status of both mothers and fathers of the conscripts, including information on the highest attained lifetime education and the annual disposable income standardised by birth years into highest-achieved quintiles between ages 40 and 50 (used to capture working life income, henceforth referred to as income categories). When values for both the mother and father were available, the highest value was retained.

### 2.7 Statistical analysis

Participants were followed from the date of conscription until the date of depression diagnosis or dispensation of antidepressants (respectively, depending on the outcome under study), emigration, death, or end of follow-up (31 December 2023), whichever came first, using age as the underlying time scale. The associations were modelled using flexible parametric survival models with baseline knots placed at the 5^th^, 27.5^th^, 50^th^, 72.5^th^, and 95^th^ percentile of the uncensored log survival times.[29, 30] We modelled cardiorespiratory fitness both as deciles and using restricted cubic splines with knots placed at the 5^th^, 35^th^, 65^th^, and 95^th^ percentile.[30] In contrast to Cox regression, the flexible parametric model directly estimates the baseline hazard, thereby enabling the computation of both hazard ratios (HRs) and absolute risk measures.[29] This enabled us to also computed the standardised cumulative incidences (1-Survival) at 65 years of age. Additionally, we estimated the preventable fraction of depression and antidepressants associated with a set of hypothetical interventions of population-levels of fitness, including a “moderate” (shifting everyone in the bottom four deciles to the fifth decile) and and “extreme” (shifting everyone to the tenth decile) intervention.[31] We performed an unadjusted model followed by a model adjusted for age at conscription (continuous), year of conscription (1972, 1973-1977, 1978-1982, 1983-1987, 1988-1992, and 1993-1995), BMI (continuous and quadratic term), IQ (continuous and quadratic), parental education (compulsory school ≤9 years, secondary education, post-secondary education <3 years, post-secondary education ≥3 years), and parental income (5 categories) (supplemental figure 2).

In exploratory analyses we examined whether BMI acted as an effect modifier by incorporating product terms between BMI categories (underweight, normal weight, overweight, obesity) and fitness deciles into the aforementioned model. We then computed marginal HRs across BMI categories for the total population while allowing for effect modification, and we post-estimated HRs within strata of BMI categories. Further, we examined the associations between fitness and the most common subtypes of depression diagnosis during follow-up, including mild (F32.0), moderate (F32.1), severe without psychotic features (F32.2), and unspecified (F32.9).

#### 2.7.1 Sibling-comparison analysis

For the sibling-comparison analysis, we extended the aforementioned model to a marginalised between-within model with robust (sandwich) standard errors, which enabled further control for unobserved shared confounders (including shared environmental and behavioural factors, and 50% genetic factors [i.e., the proportion shared between full siblings]).[18, 32] The between-within model isolates the individual-level variation (within effect) from the family-level variation (between effect) by including a term for the exposure/covariate and a term for its family average.[18, 32] All analyses were performed using Stata MP version 16.1.

#### 2.7.2 Sensitivity analyses

We performed a series of sensitivity analyses. First, to test whether differences in estimates in sibling analysis as compared to cohort analysis were more likely to be due to selection bias into the ‘sibling cohort’ rather than control for unobserved shared confounders, the standard analysis (i.e., not controlling for shared confounders) was replicated in the sibling cohort.[18] Second, because ICD-10 was not implemented until 1997, meaning that conscripts from the earlier cohorts could have been followed for many years before having the possibility to become diagnosed with depression, we repeated the analyses restricted to those conscripted in year 1985 or later (N=439 888 in the full cohort and N=119 322 full siblings). Third, to relax the proportional hazards assumption, we repeated the analyses and computed the standardised incidences at age 65 after allowing the effect of fitness to vary across time, using an interaction between a restricted cubic spline with three degress of freedom of the follow-up time (centile 33 and 67 of the distribution of the uncensored log survival times) and the fitness deciles.[29] Fourth, we repeated the analyses after ex-pressing Wmax relative to body weight (Wmax/kg), and after estimating VO_2_max using a validated equation.[33]

## 3. Results

### 3.1 Baseline characteristics

The characteristics of the 1 013 885 young men in the full cohort were similar to the 410 198 siblings (table 1). The mean age at conscription was 18.3 years, over 80% were normal weight, one in four had parents with a high (post-secondary) level of education, and one in three had parents with a high (top category) level of annual income. Compared to those with the lowest fitness, those with higher fitness were on average born slightly later, had a slightly higher BMI and IQ, and had parents with a slightly higher level of education and income (supplemental table 1).

**Table 1.** Baseline characteristics in the full cohort and the sibling cohort.

|  | **Full cohort**<br>**(N=1 013 885)** | **Sibling cohort**<br>**(N=410 198)** |
| --- | --- | --- |
| **Birth year, median (IQR)** | 1965 (1959 to 1970) | 1965 (1959 to 1969) |
| **Age at conscription, mean (SD)** | 18.3 (0.7) | 18.3 (0.6) |
| **Body mass index, kg/m<sup>2</sup>, mean (SD)** | 21.7 (2.8) | 21.7 (2.8) |
| **Body mass index categories, n (%)** |  |  |
| Underweight (<18.5 kg/m<sup>2</sup>) | 81 745 (8.1) | 33 275 (8.1) |
| Normal weight (18.5-24.9 kg/m<sup>2</sup>) | 824 468 (81.3) | 335 759 (81.9) |
| Overweight (25.0-29.9 kg/m<sup>2</sup>) | 90 664 (8.9) | 34 844 (8.5) |
| Obesity (>30.0 kg/m<sup>2</sup>) | 17 008 (1.7) | 6320 (1.5) |
| **IQ, mean (SD)** | 100 (15) | 100 (15) |
| **Cardiorespiratory fitness, Wmax, mean (SD)** | 274 (52) | 274 (52) |
| **Wmax by deciles, median (range)** |  |  |
| Decile 1 | 202 (100 to 211) | 203 (100 to 211) |
| Decile 2 | 222 (212 to 229) | 223 (212 to 229) |
| Decile 3 | 236 (230 to 241) | 236 (230 to 241) |
| Decile 4 | 250 (242 to 254) | 250 (242 to 254) |
| Decile 5 | 262 (255 to 270) | 262 (255 to 270) |
| Decile 6 | 277 (271 to 283) | 277 (271 to 282) |
| Decile 7 | 291 (284 to 300) | 291 (283 to 300) |
| Decile 8 | 311 (301 to 320) | 310 (301 to 319) |
| Decile 9 | 332 (321 to 343) | 330 (320 to 342) |
| Decile 10 | 365 (344 to 800) | 364 (343 to 800) |
| **Parental level of education, n (%)** |  |  |
| Compulsory school <9 years | 300 122 (29.6) | 125 749 (30.7) |
| Secondary education | 453 851 (44.8) | 179 448 (43.8) |
| Post-secondary education <3 years | 108 072 (10.7) | 41 318 (10.1) |
| Post-secondary education >3 years | 151 840 (15.0) | 63 683 (15.5) |
| **Parental highest income, n (%)** |  |  |
| Category 1 (low income) | 49 992 (4.9) | 16 348 (4.0) |
| Category 2 | 97 735 (9.6) | 35 796 (8.7) |
| Category 3 | 215 920 (21.3) | 87 987 (21.5) |
| Category 4 | 307 085 (30.3) | 126 230 (30.8) |
| Category 5 (high income) | 343 153 (33.9) | 143 837 (35.1) |

IQR = interquartile range. SD = standard deviation.

### 3.2 Depression and antidepressants during follow-up

Participants were followed until a median age (interquartile range [IQR]) of 56.8 (17.7 to 73.5) years, during which 47 283 (4.7%) were diagnosed with depression and 237 409 (23.4%) were dispensed antidepressants (18 150 [4.4%] and 93 150 [22.7%] respectively among siblings) (supplemental table 2, supplemental table 3). The median age at first depression diagnosis was 45.5 (39.4 to 51.5) years in the full cohort and 45.9 (40.to to 51.8) among siblings. The median age at first dispensation of antidepressants was 47.4 (41.2 to 59.1) years in the full cohort and 47.7 (41.9 to 54.4) among siblings.

### 3.3 Adolescent cardiorespiratory fitness and incident depression and dispensation of antidepressants

In cohort analysis, higher fitness levels were associated with a lower risk of diagnosis of depression and dispensation of antidepressants in a linear dose-response fashion (figure 1 & 2, table 2, supplemental table 4). Compared to the first decile, the adjusted HR for depression in the tenth decile was 0.54 (95 % confidence interval, 0.52 to 0.57), and for antidepressants 0.63 (0.62 to 0.65). The standardised cumulative incidence of depression at age 65 was 8.7% (8.5 to 8.9) in the first decile and 4.8% (4.7 to 5.0) in the tenth decile, with a difference of −3.9% (−4.1 to −3.6). For anti-depressants, the standardised cumulative incidence at age 65 was 42.0% (41.6 to 42.3) in the first decile and 29.7% (29.3 to 30.0) in the tenth decile, with a difference of −12.3% (−12.8 to −11.8). The preventable fraction at age 65 associated with hypo-thetically shifting everyone from the lowest deciles to the middle decile was 10.8% (8.6 to 13.0) for depression and 5.8% (5.0 to 6.6) for antidepressants. The preventable fraction associated with hypothetically shifting everyone to the top decile was 29.1% (26.7 to 31.6) for depression and 17.8% (16.7 to 18.6) for antidepressants (figure 3, supplemental table 5).

**Figure 1.**
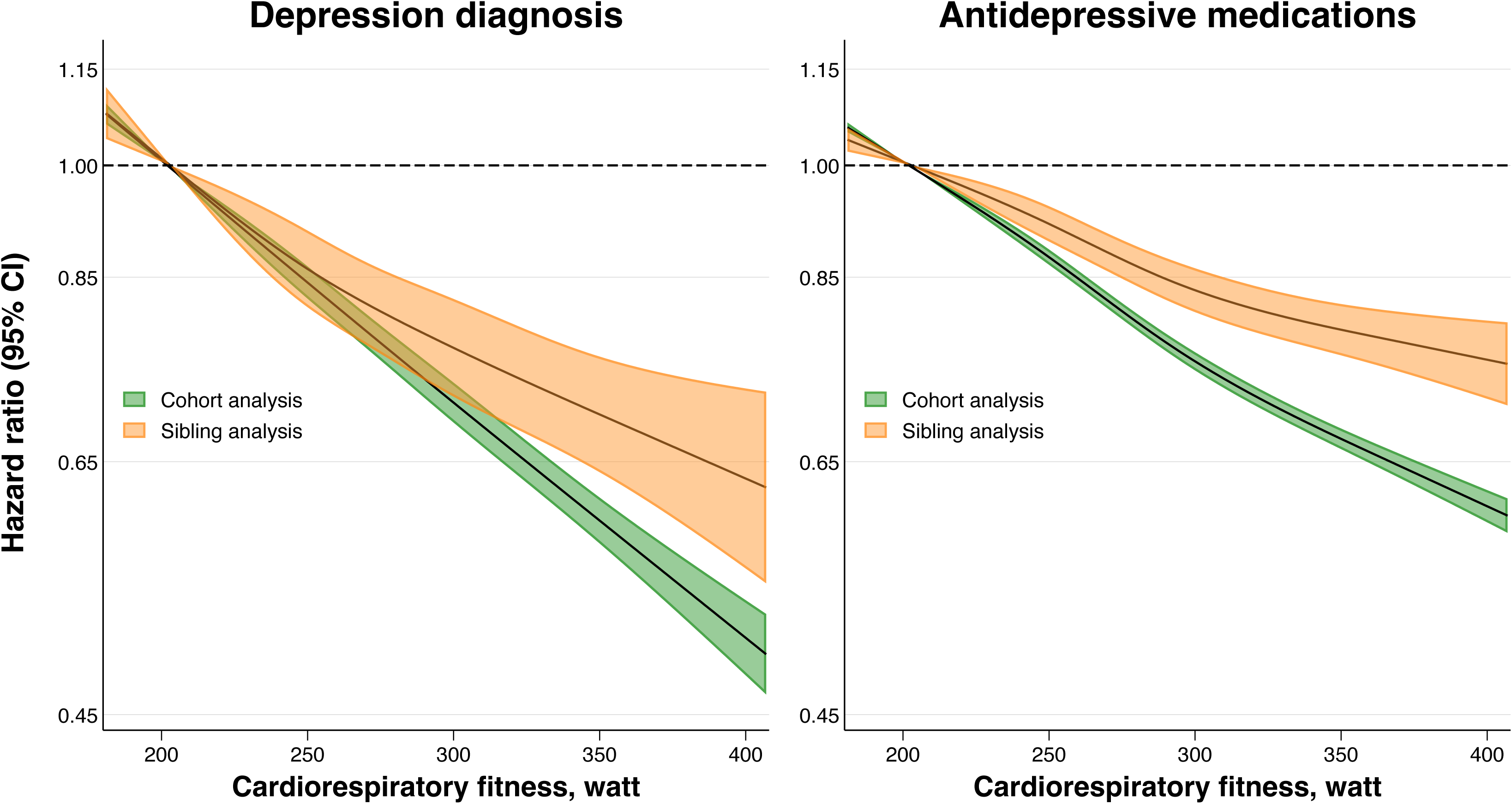
Hazard ratio for depression diagnosis and dispensation of antidepressive medications across restricted cubic splines of cardiorespiratory fitness in cohort and sibling analysis. Estimates were obtained using flexible parametric survival models, extended to a marginalised between-within model in the sibling cohort, with knots placed at the 5th, 35th, 65th, and 95th percentile, and using age as the underlying time scale. The referent was set to the median value of the bottom decile (202 Wmax). The models were adjusted for age at conscription, year of conscription, body mass index, IQ, parental education, and parental income. For graphical purposes, the x-axis was limited to span from the 1st to the 99th percentile of the exposure distribution.

**Figure 2.**
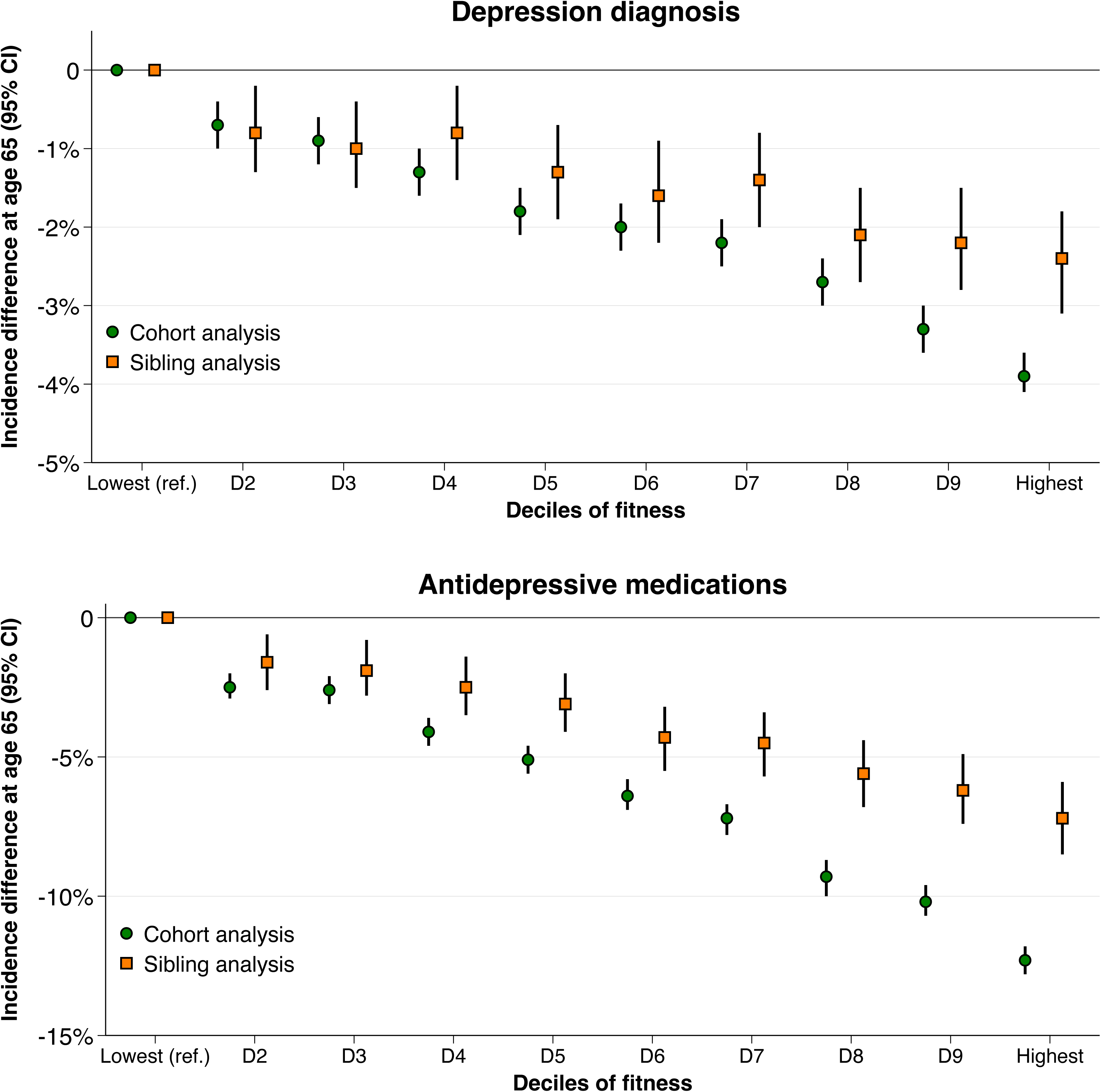
Differences in the standardised cumulative indidence of depression diagnosis and dispensation of antidepressive medications at 65 years of age by deciles of cardiorespiratory fitness in cohort and sibling analysis. Estimates were obtained using flexible parametric survival models, extended to a marginalised between-within model in the sibling cohort, with baseline knots placed at the 5^th^, 27.5^th^, 50^th^, 72.5^th^, and 95^th^ percentile, and using age as the underlying time scale. The referent was the bottom decile. The models were adjusted for age at conscription, year of conscription, body mass index, IQ, parental education, and parental income.

**Figure 3.**
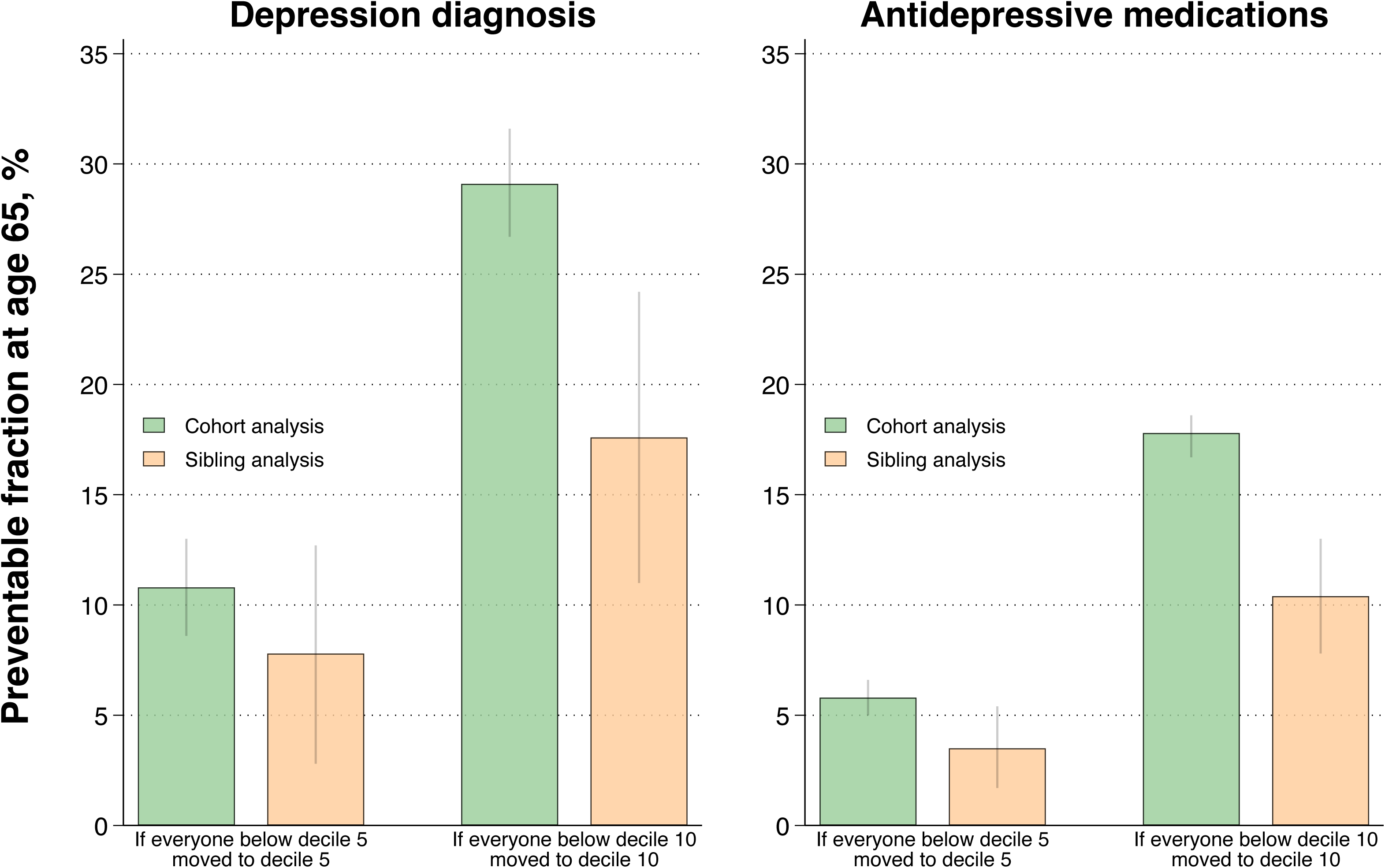
Estimated preventable fraction of depression diagnosis and antidepressive medications at 65 years of age associated with a hypothetical moderate and extreme public health intervention in cohort and sibling analysis. The extreme intervention entails shifting everyone to the top decile of cardiorespiratory fitness whereas the moderate intervention involves shifting everyone from the lowest to the middle decile of fitness. Error bars indicate 95% confidence intervals.

**Table 2.** Hazard ratios and standardised cumulative incidences of depression diagnosis and dispensation of antidepressive medications by deciles of cardiorespiratory fitness in cohort and sibling analysis.

| Depression diagnosis |  |  |  |  |  |  |  |  |
| --- | --- | --- | --- | --- | --- | --- | --- | --- |
| Deciles of fitness | Cohort analysis (N=1 013 885) |  |  |  | Sibling analysis (N=410 198) |  |  |  |
|  | Cases/N | HR (95% CI) | Standardised incidence at age 65, % (95% CI) | Incidence difference, % (95% CI) | Cases/N | HR (95% CI) | Standardised incidence at age 65, % (95% CI) | Incidence difference, % (95% CI) |
| D1 | 6905/113 887 | Ref. | 8.7 (8.5, 8.9) | Ref. | 2675/45 454 | Ref. | 7.5 (7.1, 7.9) | Ref. |
| D2 | 5392/100 857 | 0.91 (0.88, 0.95) | 8.0 (7.7, 8.2) | -0.7 (-1.0, -0.4) | 2114/41 317 | 0.89 (0.82, 0.97) | 6.7 (6.3, 7.2) | -0.8 (-1.3, -0.2) |
| D3 | 5094/96 126 | 0.89 (0.86, 0.93) | 7.8 (7.6, 8.0) | -0.9 (-1.2, -0.6) | 1975/39 087 | 0.87 (0.80, 0.94) | 6.6 (6.2, 7.0) | -1.0 (-1.5, -0.4) |
| D4 | 4985/100 065 | 0.84 (0.81, 0.88) | 7.4 (7.2, 7.6) | -1.3 (-1.6, -1.0) | 1971/41 269 | 0.89 (0.81, 0.97) | 6.7 (6.3, 7.1) | -0.8 (-1.4, -0.2) |
| D5 | 5272/110 985 | 0.79 (0.76, 0.82) | 6.9 (6.7, 7.1) | -1.8 (-2.1, -1.5) | 2016/45 169 | 0.82 (0.75, 0.90) | 6.2 (5.9, 6.6) | -1.3 (-1.9, -0.7) |
| D6 | 4019/88 231 | 0.76 (0.73, 0.79) | 6.7 (6.5, 6.9) | -2.0 (-2.3, -1.7) | 1552/36 308 | 0.79 (0.71, 0.86) | 6.0 (5.6, 6.4) | -1.6 (-2.2, -0.9) |
| D7 | 4672/103 963 | 0.74 (0.71, 0.77) | 6.5 (6.3, 6.7) | -2.2 (-2.5, -1.9) | 1724/41 950 | 0.81 (0.73, 0.89) | 6.1 (5.7, 6.5) | -1.4 (-2.0, -0.8) |
| D8 | 4164/102 896 | 0.68 (0.65, 0.71) | 6.0 (5.8, 6.2) | -2.7 (-3.0, -2.4) | 1556/41 878 | 0.71 (0.64, 0.78) | 5.4 (5.0, 5.7) | -2.1 (-2.7, -1.5) |
| D9 | 3540/96 282 | 0.61 (0.59, 0.64) | 5.4 (5.3, 5.6) | -3.3 (-3.6, -3.0) | 1307/37 917 | 0.70 (0.63, 0.78) | 5.3 (4.9, 5.7) | -2.2 (-2.8, -1.5) |
| D10 | 3240/100 593 | 0.54 (0.52, 0.57) | 4.8 (4.7, 5.0) | -3.9 (-4.1, -3.6) | 1260/39 849 | 0.67 (0.59, 0.75) | 5.1 (4.7, 5.5) | -2.4 (-3.1, -1.8) |
| Dispensation of antidepressive medications |  |  |  |  |  |  |  |  |
| Deciles of fitness | Cohort analysis (N=1 013 885) |  |  |  | Sibling analysis (N=410 198) |  |  |  |
|  | Cases/N | HR (95% CI) | Standardised incidence at age 65, % (95% CI) | Incidence difference, % (95% CI) | Cases/N | HR (95% CI) | Standardised incidence at age 65, % (95% CI) | Incidence difference, % (95% CI) |
| D1 | 32 125/113 887 | Ref. | 42.0 (41.6, 42.3) | Ref. | 12 512/45 454 | Ref. | 37.4 (36.7, 38.2) | Ref. |
| D2 | 26 175/100 857 | 0.92 (0.90, 0.93) | 39.5 (39.1, 39.9) | -2.5 (-2.9, -2.0) | 10 516/41 317 | 0.94 (0.91, 0.98) | 35.8 (35.0, 36.6) | -1.6 (-2.6, -0.6) |
| D3 | 24 919/96 126 | 0.92 (0.90, 0.93) | 39.4 (39.0, 39.8) | -2.6 (-3.1, -2.1) | 9936/39 087 | 0.93 (0.90, 0.97) | 35.5 (34.7, 36.3) | -1.9 (-2.9, -0.8) |
| D4 | 24 668/100 065 | 0.87 (0.85, 0.88) | 37.8 (37.5, 38.2) | -4.1 (-4.6, -3.6) | 9786/41 269 | 0.91 (0.88, 0.95) | 34.9 (34.2, 35.7) | -2.5 (-3.5, -1.4) |
| D5 | 26 702/110 985 | 0.84 (0.82, 0.85) | 36.9 (36.5, 37.2) | -5.1 (-5.6, -4.6) | 10 556/45 169 | 0.89 (0.86, 0.93) | 34.3 (33.6, 35.0) | -3.1 (-4.1, -2.0) |
| D6 | 20 319/88 231 | 0.80 (0.78, 0.81) | 35.6 (35.2, 36.0) | -6.4 (-6.9, -5.8) | 8081/36 308 | 0.85 (0.82, 0.89) | 33.1 (32.2, 33.8) | -4.3 (-5.5, -3.2) |
| D7 | 23 332/103 963 | 0.77 (0.76, 0.79) | 34.7 (34.4, 35.1) | -7.2 (-7.8, -6.7) | 8969/41 950 | 0.85 (0.81, 0.88) | 32.9 (32.1, 33.6) | -4.5 (-5.7, -3.4) |
| D8 | 21 379/102 896 | 0.72 (0.70, 0.73) | 32.7 (32.4, 33.1) | -9.3 (-10.0, -8.7) | 8409/41 878 | 0.81 (0.78, 0.85) | 31.8 (31.1, 32.6) | -5.6 (-6.8, -4.4) |
| D9 | 19 256/96 282 | 0.69 (0.68, 0.70) | 31.8 (31.4, 32.2) | -10.2 (-10.7, -9.6) | 7231/37 917 | 0.79 (0.76, 0.83) | 31.2 (30.4, 32.1) | -6.2 (-7.4, -4.9) |
| D10 | 18 534/100 593 | 0.63 (0.62, 0.65) | 29.7 (29.3, 30.0) | -12.3 (-12.8, -11.8) | 7154/28 775 | 0.76 (0.72, 0.80) | 30.2 (29.3, 31.1) | -7.2 (-8.5, -5.9) |

CI = confidence interval. D = decile. HR = hazard ratio. The models were adjusted for age at conscription, year of conscription, body mass index, IQ, parental education, and parental income.

Explorative analyses suggested similar estimates in the total population after allowing for interaction effects across BMI categories (supplemental table 6). When explored within strata of BMI categories, the associations seemed to be weaker in the overweight and obesity categories vs. the normal weight category (supplemental table 6), but the former was associated with lower statistical power. Further, the estimates appeared overall comparable between mild, moderate, and unspecified depression diagnosis, all of which were similar to the main outcome, whereas the associations tended to be weaker for severe depression without psychotic features (supplemental table 7).

### 3.4 Sibling-comparison analyses

When comparing full siblings, and thus controlling for all unobserved confounders shared between them, the magnitude of the associations attenuated (figure 1 & 2, table 2, supplemental table 4). Compared to the first decile of fitness, the adjusted HR in the tenth decile was 0.67 (0.59 to 0.75) for depression and 0.76 (0.72 to 0.80) for antidepressants, with incidence differences at age 65 of −2.4% (−3.1 to −1.8) and − 7.2% (−8.5 to −5.9) respectively. The preventable fraction associated with shifting everyone from the lowest deciles to the middle decile was 7.8% (2.8 to 12.7) for depression and 3.5% (1.7 to 5.4) for antidepressants, while hypothetically shifting everyone to the top decile was associated with a preventable fraction of 17.6% (11.0 to 24.2) for depression and 10.4% (7.8 to 13.0) for antidepressants (figure 3, supplemental table 5).

### 3.5 Sensitivity analyses

Replication of the standard analysis in the sibling cohort produced similar estimates as observed in the cohort analysis (supplemental table 8). Restricting the analysis to those who conscribed in year 1985 and later produced similar estimates for antidepressants, but stronger estimates for the outcome of depression diagnosis, although differences between cohort and sibling analysis remained (supplemental table 9). Allowing the effect of fitness to vary over time did not yield materially different estimates compared to the main analysis (supplemental table 10). The results were similar when using the exposures Wmax/kg and estimated VO_2_max compared to Wmax (supplemental table 11).

## 4. Discussion

In this nationwide cohort study encompassing more than 1 million men, higher levels of adolescent cardiorespiratory fitness were associated with a lower risk of adulthood depression and dispensation of antidepressants in a linear dose-response fashion. However, the associations attenuated in sibling-comparison analyses, suggesting that the findings may be influenced by familial confounding.

Our findings extend upon previous studies which have found a link between midlife fitness (typically around 40-50 years of age) and subsequent depression and antidepressant dispensation. [7, 8, 10–13] The role of specifically adolescent fitness has remained largely underexplored, although Åberg et al. reported an excess risk of adulthood depression among Swedish male conscripts with low fitness levels.[9] In our study we examined the dose-response association with even longer follow-up and greater granularity, observing lower risks already in the second decile of fitness, and the association did not seem to plateau at high levels of fitness. Our findings, regardless if proven to be causal, suggest that monitoring fitness from a young age may offer important insights into their long-term risk of mental health problems, rendering support to ongoing surveillance programs.[34, 35]

Notwithstanding this, an important question for public health and prevention is whether findings from standard observational analyses also adequately reflect a causal relationship, which would support targeted interventions, or if there are alternative explanations to the association. We found that although higher adolescent fitness were associated with lower risks of future depression and antidepressants even after controlling for unobserved confounders shared between full siblings, the magnitude of the relationships attenuated. In order to facilitate clinical interpretation of the attenuation magnitude we not only estimated HRs,[36] but we also estimated absolute cumulative risks and preventable fractions both before and after controlling for unobserved confounders shared between full siblings. Interestingly, the standardised incidence differences comparing the most fit to the least fit were notably reduced from cohort to sibling analysis; from −3.9% to −2.4% for depression, and from −12.3% to −7.2% for antidepressants. The potential public health implications of these changes are further reflected by the attenuation in the fraction of preventable cases (i.e., from 29.1% to 17.6% for depression and from 17.8% to 10.4% for antidepressants). This indicates that a sizeable proportion of the cases assumed to be preventable from improving fitness when performing standard cohort analysis may be due to familial confounding. Although the sibling-comparison analysis cannot pinpoint the definitive sources of familial confounders and their relative contribution, it may include a combination of behavioural (e.g., clustering of lifestyle habits), environmental (e.g., socioeconomic factors and upbringing), and genetic influences.

To our knowledge, no previous study has employed methods to control for familial confounding in studies on cardiorespiratory fitness and depression. Two studies did, however, incorporate twin comparators in studies on physical activity in relation to depressive and anxiety symptoms, and use of antidepressants, in predominantly middle-aged indivduals, reporting conflicting findings.[37, 38]Moreover, using a mendelian randomisation framework, Choi et al. found that genetic variants for device-measured physical activity, but not self-reported activity, were associated with a lower risk of depression.[39] Besides the difference in the exposures (physical activity vs. fitness), those estimates were unfortunately not contrasted to those derived using standard cohort analysis. Thus, our study adds new evidence by comparing standard observational estimates with those derived using sibling-comparisons, and as such demonstrating the importance of triangulation of evidence across methods to better understand the link between adolescent cardiorespiratory fitness and future risk of depressive disorders. Given the scarcity of evidence, further causal analyses are still warranted, including in studies incorporating longitudinal measures of fitness.

Another interesting finding was that the associations were weaker among individuals with a high BMI, suggesting that overweight and obesity might diminish some of the benefits associated with high fitness. While these analyses should be carefully interpreted as they were exploratory and limited in statistical power due to the small number of individuals with overweight or obesity, they may be particularly valuable for future meta-analyses because those conducted to date have not been able to examine this question.[7] [8] It would also be valuable with similar analyses based on contemporary cohorts with a higher overweight/obesity prevalence, as these might yield different results.

### 4.1 Strengths and limitations

Strengths of our study include the use and linkage of high-quality nationwide registers, allowing us to follow a large cohort of young men with standardised measures of fitness for several decades with virtually zero loss to follow-up. This provided high statistical power and enabled detailed analyses, including for sibling-comparisons. The latter also allowed for triangulation of the evidence via control for some unobserved familial confounders, in contrast to previous studies which typically relied on conventional analytical approaches.

There are also limitations. Due to historical conscription practices in Sweden, only men were included. Because the prevalence of depressive disorders is higher among women,[1] as well as evidence suggesting that sex might modify the association between fitness and depression,[12, 13] and that the genetic influence in depression may be higher in women,[40] similar studies (incorporating sibling-comparisons and/or other methods to strengthen causal inference) in women are warranted. Moreover, while we used information on dispensation of antidepressants in an attempt to capture cases in primary care, individuals who never attended specialised outpatient care but still received cognitive behavioral therapy (first line treatment) and never dispensed antidepressants are not covered by our data. Additionally, because ICD-10 was not officially implemented until 1997, and the Prescribed Drug Register was not launched until 2005, the age of first diagnosis and dispensation of medication was likely overestimated to some degree; the data is somewhat left-truncated to 1997 due to the coverage of the registries and ICD-codes. However, this is unlikely to exert an influence on the differences observed between estimates in cohort analysis compared to sibling analysis, which we also confirmed in a sensitivity analysis restricted to later cohorts. Nevertheless, the incidence might be underestimated if individuals who were diagnosed/dispensed medications prior to 1997/2005 later went into remission and none of their subsequent records noted such a history. Whether this bias would be differential or nondifferental can only be speculated. Another limitation regarding diagnoses is that we relied on registry data and were not able to clinically validate the cases. We also lacked information on certain confounders, such as loneliness, stress, alcohol and smoking, and although we compared full siblings and thus controlled for all factors that they share, residual confounding remains possible, including from non-shared behaviours, environmental factors, and genetics. Future studies based on monozygotic twins with in-depth phenotyping would therefore be valuable. Finally, while the sibling-comparison design enables control for unobserved shared factors, it hinges on assumptions including (but not limited to) the absence of non-shared confounding and no measurement error, which if present and not accounted for can amplify bias which could then explain the attenuated estimates in sibling analysis.[41]

### 4.2 Conclusion

Higher levels of adolescent cardiorespiratory fitness are associated with a lower risk of future depression diagnosis and dispensation of antidepressants, but the associations become weaker when comparing full siblings and thereby accounting for all factors shared between them. These findings highlight the challenges of obtaining valid estimates of the causal relationship between cardiorespiratory fitness and depression, and underscore the need for further causal analyses to establish more robust conclusions.

## Data Availability

The data in this study are not available to the public and will not be shared according to regulations under Swedish law. Researchers interested in obtaining the data may seek ethical approvals and inquire through Statistics Sweden. For further advice see: https://www.scb.se/en/services/guidance-for-researchers-and-universities. Analytical code underlying the results is available from the corresponding author upon request.

## Statements and declarations

### Author Contributions

MB and PN conceived the study. All authors designed the study. PN acquired the ethical permission and the data. MB performed the statistical analyses and drafted the manuscript. PN and VHA performed data managing. PN verified the underlying data. All authors interpreted the data. All authors critically revised the manuscript for intellectual content. The corresponding author MB attests that all listed authors meet authorship criteria and that no others meeting the criteria have been omitted. MB is the manuscript’s guarantors and accept full responsibility for the conduct of the study, had access to the data, and controlled the decision to publish. All authors reviewed and approved the manuscript for submission.

### Ethics approval

The study was approved by the Regional Ethical Review Board in Umeå and later by the Swedish Ethical Review Authority (no. 2010-113-31M), who waived the need for informed consent.[20]

