## Supplemental material for "Adolescent Cardiorespiratory Fitness and Future Risk of Depression and Antidepressants: A Nationwide Cohort Study with Sibling Comparators"

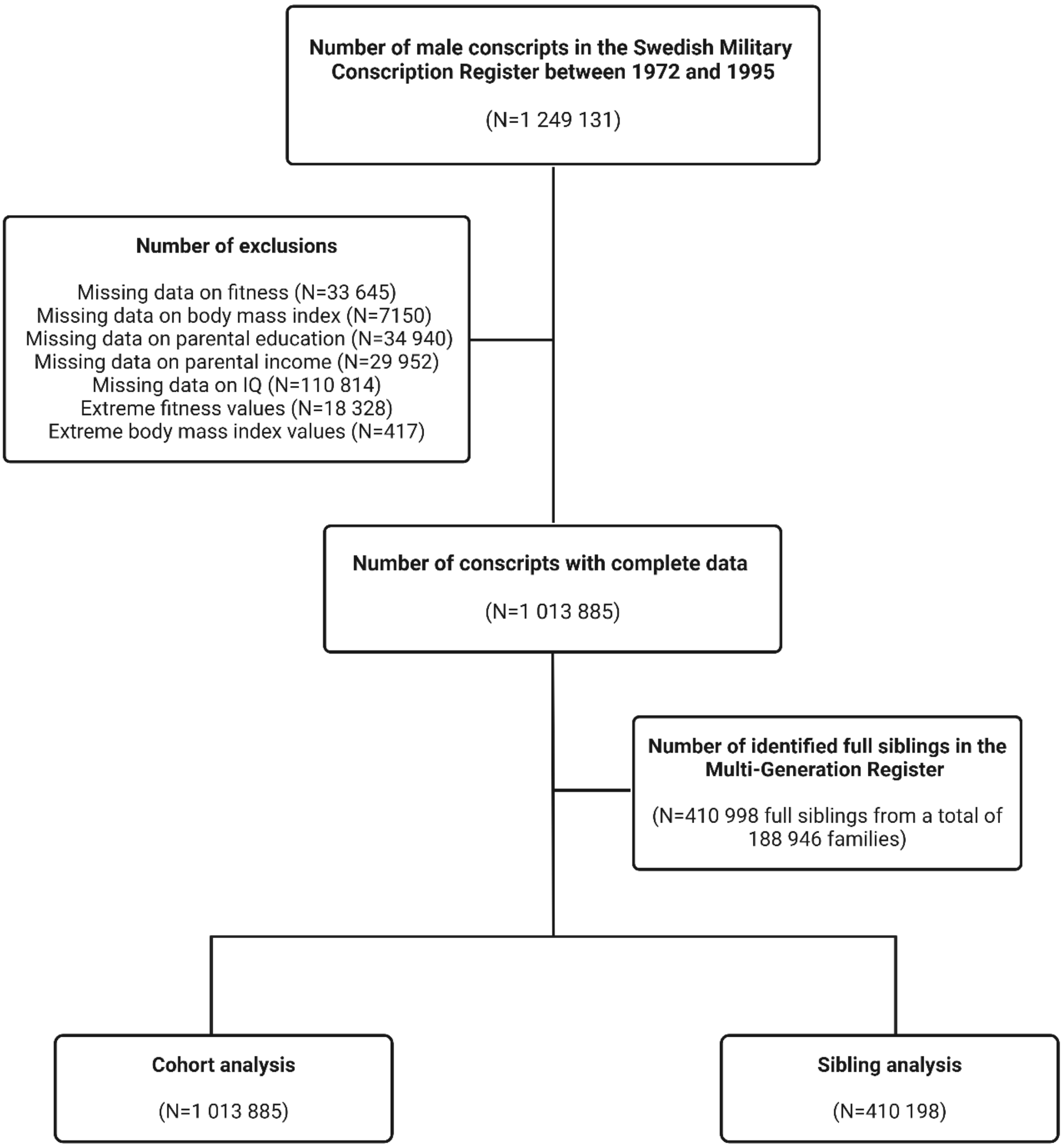


**Supplemental figure 1. Participant flow chart.** Created with BioRender.com

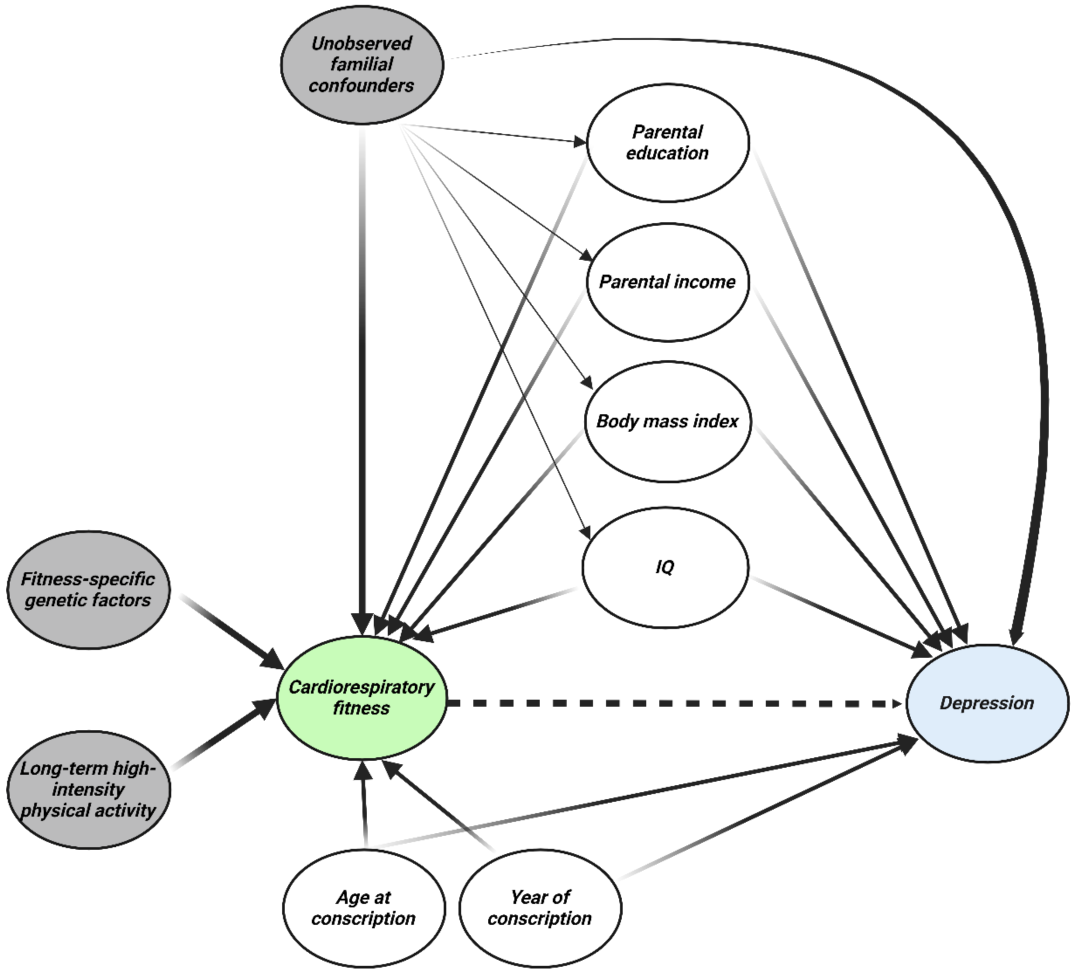


**Supplemental figure 2. Directed acyclic graph for the association between adolescent cardiorespiratory fitness and risk of depression in late adulthood, where observed (white) and unobserved (gray) confounders are illustrated.** Created with BioRender.com

### Supplemental table 1. Baseline characteristics by deciles of cardiorespiratory fitness in the full cohort and in the sibling cohort.

| | Full cohort | | | | | | | | | |
|---|---|---|---|---|---|---|---|---|---|---|
| **Variables** | **Decile 1 (N=113 887)** | **Decile 2 (N=100 857)** | **Decile 3 (N=96 126)** | **Decile 4 (N=100 065)** | **Decile 5 (N=110 985)** | **Decile 6 N=88 231)** | **Decile 7 (N=103 963)** | **Decile 8 (N=102 896)** | **Decile 9 (N=96 282)** | **Decile 10 (N=100 593)** |
| **Birth year, median (IQR)** | 1959 (1950-1976) | 1959 (1950-1976) | 1962 (1950-1976) | 1962 (1950-1977) | 1964 (1950-1977) | 1965 (1951-1977) | 1968 (1951-1977) | 1968 (1951-1977) | 1970 (1951-1977) | 1970 (1952-1977) |
| **Age at conscription, mean (SD)** | 18.4 (0.9) | 18.4 (0.8) | 18.3 (0.8) | 18.3 (0.7) | 18.3 (0.7) | 18.3 (0.6) | 18.3 (0.6) | 18.3 (0.6) | 18.3 (0.5) | 18.3 (0.5) |
| **IQ, mean (SD)** | 95.4 (15.6) | 97.8 (15.2) | 98.4 (15.1) | 99.5 (14.9) | 99.8 (14.9) | 100.7 (14.6) | 101.3 (14.5) | 102.6 (14.2) | 103.3 (13.7) | 104.5 (13.3) |
| **Body mass index, $kg/m^2$, mean (SD)** | 20.2 (2.8) | 21.0 (2.7) | 21.4 (2.9) | 21.6 (2.8) | 21.8 (2.8) | 21.9 (2.7) | 22.1 (2.8) | 22.3 (2.6) | 22.3 (2.5) | 23.0 (2.6) |
| **Body mass index categories, n (%)** | | | | | | | | | | |
| Underweight (<18.5 $kg/m^2$) | 28 940 (25.4) | 13 490 (13.4) | 9739 (10.1) | 7802 (7.8) | 7464 (6.7) | 4605 (5.2) | 4388 (4.2) | 2604 (2.5) | 2067 (2.2) | 646 (0.6) |
| Normal weight (18.5-24.9 $kg/m^2$) | 78 788 (69.2) | 80 030 (79.4) | 77 270 (80.4) | 82 466 (82.4) | 91 688 (82.6) | 73 900 (83.8) | 86 937 (83.6) | 87 210 (84.8) | 83 007 (86.2) | 83 172 (82.7) |
| Overweight (25.0-29.9 $kg/m^2$) | 4814 (4.2) | 6046 (6.0) | 7502 (7.8) | 8107 (8.1) | 9844 (8.9) | 8213 (9.3) | 10 538 (10.1) | 11 320 (11.0) | 9718 (10.1) | 14 562 (14.5) |
| Obesity ($\geq$30.0 $kg/m^2$) | 1345 (1.2) | 1291 (1.3) | 1615 (1.7) | 1690 (1.7) | 1989 (1.8) | 1513 (1.7) | 2100 (2.0) | 1762 (1.7) | 1490 (1.6) | 2213 (2.2) |
| **Parental level of education, n (%)** | | | | | | | | | | |
| Compulsory school $\leq$9 years | 49 379 (43.4) | 41 529 (41.2) | 36 126 (37.6) | 34 806 (34.8) | 33 929 (30.6) | 25 090 (28.4) | 24 932 (24.0) | 22 524 (21.9) | 16 903 (17.6) | 14 904 (14.8) |
| Secondary education | 47 694 (41.9) | 42 669 (42.3) | 41 827 (43.5) | 44 282 (44.3) | 50 781 (45.8) | 40 414 (45.8) | 48 885 (47.0) | 47 487 (46.2) | 44 776 (46.5) | 45 036 (44.8) |
| Post-secondary education <3 years | 7737 (6.8) | 7465 (7.4) | 7974 (8.3) | 9119 (9.1) | 11 370 (10.2) | 9410 (10.7) | 12 563 (12.1) | 13 252 (12.9) | 13 802 (14.3) | 15 380 (15.3) |
| Post-secondary education $\geq$3 years | 9077 (8.0) | 9194 (9.1) | 10 199 (10.6) | 11 858 (11.9) | 14 905 (13.4) | 13 317 (15.1) | 17 583 (16.9) | 19 633 (19.1) | 20 801 (21.6) | 25 273 (25.1) |
| **Parental highest income, n (%)** | | | | | | | | | | |
| Category 1 (low income) | 7879 (6.9) | 6522 (6.5) | 5697 (5.9) | 5645 (5.6) | 5636 (5.1) | 4295 (4.8) | 4516 (4.3) | 3897 (3.8) | 3137 (3.3) | 2768 (2.8) |
| Category 2 | 13 690 (12.0) | 11 358 (11.3) | 10 445 (10.9) | 10 267 (10.3) | 11 325 (10.2) | 8231 (9.3) | 9443 (9.1) | 8570 (8.3) | 7647 (7.9) | 6759 (6.7) |
| Category 3 | 28 519 (25.0) | 24 167 (24.0) | 22 139 (23.0) | 22 436 (22.4) | 24 161 (21.8) | 18 498 (21.0) | 21 338 (20.5) | 19 739 (19.2) | 17 866 (18.6) | 17 057 (17.0) |
| Category 4 | 34 908 (30.7) | 30 443 (30.2) | 29 576 (30.8) | 30 619 (30.6) | 33 749 (30.4) | 26 847 (30.4) | 31 532 (30.3) | 30 630 (29.8) | 28 985 (30.1) | 29 796 (29.6) |
| Category 5 (high income) | 28 891 (25.4) | 28 367 (28.1) | 28 269 (29.4) | 31 098 (31.1) | 36 114 (32.5) | 30 360 (34.4) | 37 134 (35.7) | 40 060 (38.9) | 38 647 (40.1) | 44 213 (44.0) |
| | **Sibling cohort** | | | | | | | | | |
| | **Decile 1 (N=45 454)** | **Decile 2 (N=41 317)** | **Decile 3 (N=39 087)** | **Decile 4 (N=41 269)** | **Decile 5 (N=45 169)** | **Decile 6 (N=36 308)** | **Decile 7 (N=41 950)** | **Decile 8 (N=41 878)** | **Decile 9 (N=37 917)** | **Decile 10 (N=39 849)** |
| **Birth year, median (IQR)** | 1960 (1950-1976) | 1961 (1950-1976) | 1962 (1950-1976) | 1962 (1950-1976) | 1964 (1951-1977) | 1965 (1951-1976) | 1967 (1951-1977) | 1967 (1952-1977) | 1969 (1952-1977) | 1970 (1953-1977) |
| **Age at conscription, mean (SD)** | 18.4 (0.9) | 18.3 (0.8) | 18.3 (0.8) | 18.3 (0.7) | 18.3 (0.7) | 18.3 (0.6) | 18.3 (0.6) | 18.2 (0.5) | 18.3 (0.5) | 18.3 (0.5) |
| **IQ, mean (SD)** | 94.6 (15.6) | 97.1 (15.3) | 97.8 (15.3) | 99.0 (15.1) | 99.6 (15.0) | 100.6 (14.7) | 101.2 (14.6) | 102.6 (14.3) | 103.2 (13.8) | 104.7 (13.4) |
| **Body mass index, $kg/m^2$, mean (SD)** | 20.2 (2.7) | 20.9 (2.7) | 21.4 (2.8) | 21.5 (2.8) | 21.7 (2.8) | 21.8 (2.7) | 22.0 (2.8) | 22.3 (2.6) | 22.2 (2.5) | 22.9 (2.6) |
| **Body mass index categories, n (%)** | | | | | | | | | | |
| Underweight (<18.5 $kg/m^2$) | 11 711 (25.8) | 5646 (13.7) | 3941 (10.1) | 3167 (7.7) | 3009 (6.7) | 1868 (5.1) | 1742 (4.2) | 1078 (2.6) | 840 (2.2) | 273 (0.7) |
| Normal weight (18.5-24.9 $kg/m^2$) | 31 449 (69.2) | 32 801 (79.4) | 31 550 (80.7) | 34 267 (83.0) | 37 575 (83.2) | 30 678 (84.5) | 35 394 (84.4) | 35 772 (85.4) | 32 875 (86.7) | 33 398 (83.8) |
| Overweight (25.0-29.9 $kg/m^2$) | 1824 (4.0) | 2376(5.8) | 2973 (7.6) | 3176 (7.7) | 3827 (8.5) | 3210 (8.8) | 4036 (9.6) | 4366 (10.4) | 3661 (9.7) | 5395 (13.5) |
| Obesity ($\geq$30.0 $kg/m^2$) | 470 (1.0) | 494 (1.2) | 623 (1.6) | 659 (1.6) | 758 (1.7) | 552 (1.5) | 778 (1.9) | 662 (1.6) | 541 (1.4) | 783 (2.0) |
| **Parental level of education, n (%)** | | | | | | | | | | |
| Compulsory school $\leq$9 years | 20 026 (44.1) | 17 083 (41.4) | 14 863 (38.0) | 14 579 (35.3) | 14 426 (31.9) | 10 761 (29.6) | 10 707 (25.5) | 9751 (23.3) | 7251 (19.1) | 6302 (15.8) |
| Secondary education | 18 756 (41.3) | 17 285 (41.8) | 16 845 (43.1) | 17 900 (43.4) | 20 141 (44.6) | 16 110 (44.4) | 19 218 (45.8) | 18 670 (44.6) | 17 198 (45.4) | 17 325 (43.5) |
| Post-secondary education <3 years | 2965 (6.5) | 2943 (7.1) | 3050 (7.8) | 3607 (8.7) | 4396 (9.7) | 3714 (10.2) | 4746 (11.3) | 5069 (12.1) | 5074 (13.4) | 5754 (14.4) |
| Post-secondary education $\geq$3 years | 3707 (8.2) | 4006 (9.7) | 4329 (11.1) | 5183 (12.6) | 6206 (13.7) | 5723 (15.8) | 7279 (17.4) | 8388 (20.0) | 8394 (22.1) | 10 468 (26.3) |
| **Parental highest income, n (%)** | | | | | | | | | | |
| Category 1 (low income) | 2525 (5.6) | 2152 (5.2) | 1834 (4.7) | 1837 (4.5) | 1830 (4.1) | 1429 (3.9) | 1497 (3.6) | 1289 (3.1) | 1047 (2.8) | 908 (2.3) |
| Category 2 | 5026 (11.1) | 4286 (10.4) | 3918 (10.0) | 3774 (9.1) | 4179 (9.3) | 3050 (8.4) | 3445 (8.2) | 3094 (7.4) | 2619 (6.9) | 2405 (6.0) |
| Category 3 | 11 782 (25.9) | 10 090 (24.4) | 9086 (23.3) | 9359 (22.7) | 10 032 (22.2) | 7564 (20.8) | 8613 (20.5) | 7890 (18.8) | 6899 (18.2) | 6672 (16.7) |
| Category 4 | 14 520 (31.9) | 12 762 (30.9) | 12 332 (31.6) | 12 914 (31.3) | 13 947 (30.9) | 11 247 (31.0) | 12 787 (30.5) | 12 586 (30.1) | 11 477 (30.3) | 11 658 (29.3) |
| Category 5 (high income) | 11 601 (25.5) | 12 027 (29.1) | 11 917 (30.5) | 13 385 (32.4) | 15 181 (33.6) | 13 018 (35.9) | 15 608 (37.2) | 17 019 (40.6) | 15 875 (41.9) | 18 206 (45.7) |

IQR = interquartile range. SD = standard deviation.

### Supplemental table 2. Numbers censored due, death, emigration, and end of follow-up.

| | Cohort analysis (N=1 013 885) | Sibling analysis (N=410 998) |
|---|---|---|
| **Depression diagnosis** | | |
| Outcome | 47 283 (4.7) | 18 150 (4.4) |
| Death | 55 409 (5.5) | 21 227 (5.2) |
| Emigration | 65 043 (6.4) | 25 457 (6.2) |
| End of follow-up | 846 150 (83.5) | 345 364 (84.2) |
| **Antidepressive medications** | | |
| Outcome | 237 409 (23.4) | 93 150 (22.7) |
| Death | 43 380 (4.3) | 16 647 (4.1) |
| Emigration | 56 448 (5.6) | 22 237 (5.4) |
| End of follow-up | 676 648 (66.7) | 278 164 (67.8) |

Number of events and numbers censored are shown as n (%).

### Supplemental table 3. Subtypes of depression diagnoses among individuals with a depression outcome during follow-up.

| Depression subtype, ICD-10 code | N cases in the full cohort | N cases in the sibling cohort |
|---|---|---|
| Major depressive disorder, single episode, mild, F32.0 | 4801 | 1803 |
| Major depressive disorder, single episode, moderate, F32.1 | 11 570 | 4379 |
| Major depressive disorder, single episode, severe without psychotic features, F32.2 | 4697 | 1920 |
| Major depressive disorder, single episode, severe with psychotic features, F32.3 | 1271 | 531 |
| Major depressive disorder, single episode, in partial remission, F32.4 | 1 | 0 |
| Other depressive episodes, F32.8 | 615 | 253 |
| Major depressive disorder, single episode, unspecified, F32.9 | 24 444 | 9308 |

### Supplemental table 4. Unadjusted hazard ratios for depression diagnosis and dispensation of antidepressive medications by deciles of cardiorespiratory fitness in the full cohort.

| | Depression diagnosis | | Dispensation of antidepressive medications | |
|---|---|---|---|---|
| Deciles of fitness | Cases/N | HR (95% CI) | Cases/N | HR (95% CI) |
| D1 | 6905/113 887 | Ref. | 32 125/113 887 | Ref. |
| D2 | 5392/100 857 | 0.87 (0.84, 0.90) | 26 175/100 857 | 0.90 (0.88, 0.91) |
| D3 | 5094/96 126 | 0.90 (0.87, 0.93) | 24 919/96 126 | 0.95 (0.94, 0.97) |
| D4 | 4985/100 065 | 0.87 (0.83, 0.90) | 24 668/100 065 | 0.93 (0.91, 0.95) |
| D5 | 5272/110 985 | 0.88 (0.85, 0.91) | 26 702/110 985 | 0.99 (0.97, 1.01) |
| D6 | 4019/88 231 | 0.85 (0.82, 0.89) | 20 319/88 231 | 0.96 (0.94, 0.98) |
| D7 | 4672/103 963 | 0.90 (0.87, 0.94) | 23 332/103 963 | 1.04 (1.02, 1.06) |
| D8 | 4164/102 896 | 0.81 (0.78, 0.85) | 21 379/102 896 | 0.96 (0.94, 0.98) |
| D9 | 3540/96 282 | 0.81 (0.78, 0.84) | 19 256/96 282 | 1.05 (1.03, 1.07) |
| D10 | 3240/100 593 | 0.72 (0.69, 0.76) | 18 534/100 593 | 0.99 (0.97, 1.01) |

D = decile. CI = confidence interval. HR = hazard ratio.

**Supplemental table 5. Estimated preventable fraction of depression diagnosis and dispensation of antidepressive medications at 65 years of age associated with a moderate (shifting those below deciles 5 to decile 5), or an extreme hypothetical intervention (shifting everyone to decile 10), in cohort and sibling analysis.**

| | Depression diagnosis | |
|---|---|---|
| | **Cohort analysis (N=1 013 885)** | **Sibling analysis (N=410 198)** |
| **Hypothetical intervention** | **Preventable fraction, % (95% CI)** | **Preventable fraction, % (95% CI)** |
| Moderate | 10.8 (8.6, 13.0) | 7.8 (2.8 to 12.7) |
| Extreme | 29.1 (26.7, 31.6) | 17.6 (11.0, 24.2) |

| | Dispensation of antidepressive medications | |
|---|---|---|
| | **Cohort analysis (N=1 013 885)** | **Sibling analysis (N=410 198)** |
| **Hypothetical intervention** | **Preventable fraction, % (95% CI)** | **Preventable fraction, % (95% CI)** |
| Moderate | 5.8 (5.0, 6.6) | 3.5 (1.7, 5.4) |
| Extreme | 17.8 (16.7, 18.6) | 10.4 (7.8, 13.0) |

CI = confidence interval. The models were adjusted for age at conscription, year of conscription, body mass index, IQ, parental education, and parental income.

**Supplemental table 6. Hazard ratios for depression diagnosis and dispensation of antidepressive medications by deciles of cardiorespiratory fitness with and without allowing for effect modification by BMI, in cohort and sibling analysis.**

| | Fully adjusted model assuming no effect modification by BMI (as reported in the main article)[a] | | | Fully adjusted model incorporating interaction terms between fitness and BMI[b] | |
|---|---|---|---|---|---|
| | **Cohort analysis (N=1 013 885)** | **Sibling analysis (N=410 198)** | | **Cohort analysis (N=1 013 885** | **Sibling analysis (N=410 198)** |
| **Deciles of fitness** | **HR (95% CI)** | **HR (95% CI)** | **Deciles of fitness** | **HR (95% CI)** | **HR (95% CI)** |
| | **Depression diagnosis** | | | **Depression diagnosis** | |
| **Total population** | | | **Total population** | | |
| D1 | Ref. | Ref. | D1 | Ref. | Ref. |
| D2 | 0.91 (0.88, 0.95) | 0.89 (0.82, 0.97) | D2 | 0.90 (0.87, 0.93) | 0.86 (0.79, 0.94) |
| D3 | 0.89 (0.86, 0.93) | 0.87 (0.80, 0.94) | D3 | 0.88 (0.85, 0.92) | 0.87 (0.80, 0.95) |
| D4 | 0.84 (0.81, 0.88) | 0.89 (0.81, 0.97) | D4 | 0.84 (0.81, 0.87) | 0.87 (0.80, 0.96) |
| D5 | 0.79 (0.76, 0.82) | 0.82 (0.75, 0.90) | D5 | 0.77 (0.74, 0.80) | 0.81 (0.74, 0.88) |
| D6 | 0.76 (0.73, 0.79) | 0.79 (0.71, 0.86) | D6 | 0.75 (0.72, 0.79) | 0.77 (0.70, 0.85) |
| D7 | 0.74 (0.71, 0.77) | 0.81 (0.73, 0.89) | D7 | 0.73 (0.70, 0.76) | 0.79 (0.72, 0.87) |
| D8 | 0.68 (0.65, 0.71) | 0.71 (0.64, 0.78) | D8 | 0.66 (0.64, 0.69) | 0.68 (0.62, 0.75) |
| D9 | 0.61 (0.59, 0.64) | 0.70 (0.63, 0.78) | D9 | 0.60 (0.57, 0.63) | 0.68 (0.61, 0.75) |
| D10 | 0.54 (0.52, 0.57) | 0.67 (0.59, 0.75) | D10 | 0.53 (0.50, 0.55) | 0.64 (0.57, 0.72) |
| | | | **Underweight** | | |
| | | | D1 | Ref. | Ref. |
| | | | D2 | 0.96 (0.89, 1.03) | 1.07 (0.90, 1.26) |
| | | | D3 | 0.94 (0.86, 1.02) | 0.77 (0.64, 0.94) |
| | | | D4 | 0.93 (0.85, 1.03) | 0.96 (0.77, 1.19) |
| | | | D5 | 0.80 (0.72, 0.89) | 0.81 (0.64, 1.03) |
| | | | D6 | 0.79 (0.69, 0.90) | 0.84 (0.62, 1.14) |
| | | | D7 | 0.70 (0.60, 0.81) | 0.98 (0.70, 1.38) |
| | | | D8 | 0.73 (0.60, 0.88) | 0.73 (0.48, 1.10) |
| | | | D9 | 0.53 (0.41, 0.67) | 0.68 (0.39, 1.16) |
| | | | D10 | 0.52 (0.33, 0.82) | 0.54 (0.24, 1.21) |
| | | | **Normal weight** | | |
| | | | D1 | Ref. | Ref. |
| | | | D2 | 0.90 (0.87, 0.93) | 0.86 (0.79, 0.94) |
| | | | D3 | 0.88 (0.85, 0.92) | 0.87 (0.80, 0.95) |
| | | | D4 | 0.84 (0.81, 0.87) | 0.87 (0.80, 0.96) |
| | | | D5 | 0.77 (0.74, 0.80) | 0.81 (0.74, 0.88) |
| | | | D6 | 0.75 (0.72, 0.79) | 0.77 (0.70, 0.85) |
| | | | D7 | 0.73 (0.70, 0.76) | 0.79 (0.72, 0.87) |
| | | | D8 | 0.66 (0.64, 0.69) | 0.68 (0.62, 0.75) |
| | | | D9 | 0.60 (0.57, 0.63) | 0.68 (0.61, 0.75) |
| | | | D10 | 0.53 (0.50, 0.55) | 0.65 (0.57, 0.72) |
| | | | **Overweight** | | |
| | | | D1 | Ref. | Ref. |
| | | | D2 | 0.96 (0.86, 1.06) | 0.91 (0.71, 1.16) |
| | | | D3 | 0.90 (0.81, 0.99) | 0.89 (0.72, 1.10) |
| | | | D4 | 0.84 (0.76, 0.93) | 0.90 (0.72, 1.13) |
| | | | D5 | 0.88 (0.81, 0.96) | 0.78 (0.64, 0.95) |
| | | | D6 | 0.80 (0.73, 0.88) | 0.83 (0.67, 1.03) |
| | | | D7 | 0.83 (0.76, 0.90) | 0.79 (0.64, 0.98) |
| | | | D8 | 0.75 (0.69, 0.82) | 0.82 (0.67, 1.01) |
| | | | D9 | 0.67 (0.61, 0.74) | 0.72 (0.57, 0.90) |
| | | | D10 | 0.61 (0.56, 0.67) | 0.74 (0.60, 0.91) |
| | | | **Obesity** | | |
| | | | D1 | Ref. | Ref. |
| | | | D2 | 0.91 (0.73, 1.14) | 0.79 (0.48, 1.27) |
| | | | D3 | 1.05 (0.87, 1.26) | 1.05 (0.69, 1.62) |
| | | | D4 | 0.77 (0.62, 0.95) | 0.68 (0.41, 1.14) |
| | | | D5 | 1.06 (0.90, 1.26) | 1.42 (0.94, 2.15) |
| | | | D6 | 0.79 (0.64, 0.99) | 0.64 (0.39, 1.07) |
| | | | D7 | 0.69 (0.57, 0.84) | 0.71 (0.46, 1.09) |
| | | | D8 | 0.83 (0.68, 1.02) | 0.80 (0.51, 1.25) |
| | | | D9 | 0.96 (0.79, 1.18) | 1.60 (0.88, 2.92) |
| | | | D10 | 0.74 (0.61, 0.89) | 0.65 (0.44, 0.97) |

| | Dispensation of antidepressive medications | |
|---|---|---|
| **Total population** | | |
| D1 | Ref. | Ref. |
| D2 | 0.92 (0.90, 0.93) | 0.94 (0.91, 0.98) |
| D3 | 0.92 (0.90, 0.93) | 0.93 (0.90, 0.97) |
| D4 | 0.87 (0.85, 0.88) | 0.91 (0.88, 0.95) |
| D5 | 0.84 (0.82, 0.85) | 0.89 (0.86, 0.93) |
| D6 | 0.80 (0.78, 0.81) | 0.85 (0.82, 0.89) |
| D7 | 0.77 (0.76, 0.79) | 0.85 (0.81, 0.88) |
| D8 | 0.72 (0.70, 0.73) | 0.81 (0.78, 0.85) |
| D9 | 0.69 (0.68, 0.70) | 0.79 (0.76, 0.83) |
| D10 | 0.63 (0.62, 0.65) | 0.76 (0.72, 0.80) |

| | Dispensation of antidepressive medications | |
|---|---|---|
| **Total population** | | |
| D1 | Ref. | Ref. |
| D2 | 0.92 (0.91, 0.94) | 0.94 (0.91, 0.97) |
| D3 | 0.92 (0.91, 0.94) | 0.94 (0.90, 0.97) |
| D4 | 0.88 (0.86, 0.89) | 0.91 (0.88, 0.95) |
| D5 | 0.85 (0.83, 0.86) | 0.90 (0.86, 0.93) |
| D6 | 0.82 (0.80, 0.83) | 0.85 (0.82, 0.89) |
| D7 | 0.79 (0.78, 0.80) | 0.85 (0.81, 0.89) |
| D8 | 0.73 (0.72, 0.75) | 0.82 (0.79, 0.86) |
| D9 | 0.71 (0.70, 0.72) | 0.80 (0.77, 0.84) |
| D10 | 0.66 (0.64, 0.67) | 0.76 (0.73, 0.80) |
| **Underweight** | | |
| D1 | Ref. | Ref. |
| D2 | 0.94 (0.92, 0.97) | 0.97 (0.91, 1.04) |
| D3 | 0.93 (0.90, 0.97) | 0.95 (0.87, 1.03) |
| D4 | 0.94 (0.90, 0.98) | 1.02 (0.93, 1.12) |
| D5 | 0.85 (0.82, 0.89) | 0.87 (0.79, 0.97) |
| D6 | 0.78 (0.74, 0.83) | 0.87 (0.76, 0.99) |
| D7 | 0.80 (0.76, 0.85) | 0.93 (0.80, 1.07) |
| D8 | 0.75 (0.69, 0.81) | 0.79 (0.67, 0.95) |
| D9 | 0.76 (0.69, 0.83) | 0.70 (0.57, 0.87) |
| D10 | 0.69 (0.59, 0.82) | 0.74 (0.52, 1.08) |
| **Normal weight** | | |
| D1 | Ref. | Ref. |
| D2 | 0.92 (0.91, 0.94) | 0.94 (0.91, 0.97) |
| D3 | 0.92 (0.91, 0.94) | 0.94 (0.90, 0.97) |
| D4 | 0.88 (0.86, 0.89) | 0.91 (0.88, 0.95) |
| D5 | 0.85 (0.83, 0.86) | 0.90 (0.86, 0.93) |
| D6 | 0.82 (0.80, 0.83) | 0.85 (0.82, 0.89) |
| D7 | 0.79 (0.78, 0.80) | 0.85 (0.82, 0.89) |
| D8 | 0.73 (0.72, 0.75) | 0.82 (0.79, 0.86) |
| D9 | 0.71 (0.70, 0.72) | 0.80 (0.77, 0.84) |
| D10 | 0.66 (0.64, 0.67) | 0.76 (0.73, 0.80) |
| **Overweight** | | |
| D1 | Ref. | Ref. |
| D2 | 0.96 (0.92, 1.01) | 1.03 (0.93, 1.14) |
| D3 | 0.97 (0.93, 1.01) | 0.98 (0.89, 1.07) |
| D4 | 0.90 (0.87, 0.94) | 0.93 (0.85, 1.02) |
| D5 | 0.91 (0.88, 0.95) | 0.94 (0.86, 1.03) |
| D6 | 0.89 (0.86, 0.93) | 0.95 (0.87, 1.04) |
| D7 | 0.86 (0.83, 0.89) | 0.89 (0.81, 0.97) |
| D8 | 0.80 (0.78, 0.83) | 0.84 (0.77, 0.92) |
| D9 | 0.78 (0.75, 0.81) | 0.83 (0.75, 0.91) |
| D10 | 0.71 (0.68, 0.73) | 0.86 (0.79, 0.94) |
| **Obesity** | | |
| D1 | Ref. | Ref. |
| D2 | 0.94 (0.85, 1.03) | 0.99 (0.79, 1.24) |
| D3 | 0.96 (0.88, 1.04) | 0.89 (0.74, 1.09) |
| D4 | 0.95 (0.88, 1.03) | 0.86 (0.70, 1.05) |
| D5 | 1.02 (0.94, 1.09) | 1.14 (0.97, 1.35) |
| D6 | 0.91 (0.83, 0.99) | 0.85 (0.69, 1.05) |
| D7 | 0.89 (0.83, 0.96) | 0.90 (0.75, 1.09) |
| D8 | 0.89 (0.82, 0.96) | 0.88 (0.72, 1.08) |
| D9 | 0.91 (0.83, 1.00) | 1.12 (0.89, 1.40) |
| D10 | 0.83 (0.77, 0.90) | 0.82 (0.68, 1.00) |

BMI = body mass index. D = decile. CI = confidence interval. HR = hazard ratio.

[a]Adjusted for age at conscription, year of conscription, IQ, BMI, parental education, and parental income.

[b]Adjusted for age at conscription, year of conscription, IQ, BMI, parental education, parental income (and additionally for interaction terms between fitness and BMI categories in the total population).

### Supplemental table 7. Hazard ratios for depression subtypes by deciles of cardiorespiratory fitness in cohort and sibling analysis.

#### Major depressive disorder, mild (F32.0)

| Deciles of fitness | Cohort analysis (N=1 013 885) | | | Sibling analysis (N=410 198) | | |
|---|---|---|---|---|---|---|
| | Cases/N | HR | 95% CI | Cases/N | HR | 95% CI |
| D1 | 700/113 887 | Ref. | - | 282/45 454 | Ref. | - |
| D2 | 569/100 857 | 0.95 | 0.85, 1.06 | 222/41 317 | 0.82 | 0.64, 1.06 |
| D3 | 498/96 126 | 0.86 | 0.76, 0.96 | 188/39 087 | 0.74 | 0.57, 0.97 |
| D4 | 502/100 065 | 0.83 | 0.74, 0.93 | 190/41 269 | 0.76 | 0.58, 0.99 |
| D5 | 501/110 985 | 0.72 | 0.64, 0.81 | 199/45 169 | 0.75 | 0.57, 0.98 |
| D6 | 415/88 231 | 0.76 | 0.66, 0.86 | 154/36 308 | 0.62 | 0.46, 0.84 |
| D7 | 454/103 963 | 0.68 | 0.60, 0.77 | 165/41 950 | 0.72 | 0.53, 0.97 |
| D8 | 435/102 896 | 0.67 | 0.59, 0.77 | 158/41 878 | 0.57 | 0.41, 0.77 |
| D9 | 397/96 282 | 0.64 | 0.56, 0.74 | 133/37 917 | 0.55 | 0.39, 0.76 |
| D10 | 330/100 593 | 0.52 | 0.45, 0.60 | 112/39 849 | 0.46 | 0.32, 0.66 |

#### Major depressive disorder, moderate (F32.1)

| Deciles of fitness | Cohort analysis (N=1 013 885) | | | Sibling analysis (N=410 198) | | |
|---|---|---|---|---|---|---|
| | Cases/N | HR | 95% CI | Cases/N | HR | 95% CI |
| D1 | 1589/113 887 | Ref. | - | 611/45 454 | Ref. | - |
| D2 | 1273/100 857 | 0.93 | 0.87, 1.00 | 475/41 317 | 0.91 | 0.77, 1.08 |
| D3 | 1144/96 126 | 0.85 | 0.79, 0.92 | 440/39 087 | 0.78 | 0.65, 0.92 |
| D4 | 1221/100 065 | 0.87 | 0.81, 0.94 | 475/41 269 | 0.86 | 0.72, 1.03 |
| D5 | 1348/110 985 | 0.83 | 0.77, 0.89 | 515/45 169 | 0.87 | 0.73, 1.04 |
| D6 | 994/88 231 | 0.77 | 0.71, 0.84 | 376/36 308 | 0.82 | 0.68, 1.00 |
| D7 | 1249/103 963 | 0.79 | 0.73, 0.86 | 457/41 950 | 0.82 | 0.68, 1.00 |
| D8 | 1041/102 896 | 0.68 | 0.62, 0.74 | 383/41 878 | 0.71 | 0.58, 0.87 |
| D9 | 868/96 282 | 0.59 | 0.54, 0.64 | 332/37 917 | 0.65 | 0.53, 0.81 |
| D10 | 843/100 593 | 0.55 | 0.50, 0.60 | 315/39 849 | 0.65 | 0.51, 0.82 |

#### Major depressive disorder, severe without psychotic features (F32.2)

| Deciles of fitness | Cohort analysis (N=1 013 885) | | | Sibling analysis (N=410 198) | | |
|---|---|---|---|---|---|---|
| | Cases/N | HR | 95% CI | Cases/N | HR | 95% CI |
| D1 | 689/113 887 | Ref. | - | 278/45 454 | Ref. | - |
| D2 | 532/100 857 | 0.91 | 0.81, 1.02 | 239/41 317 | 0.96 | 0.74, 1.23 |
| D3 | 515/96 126 | 0.92 | 0.82, 1.04 | 200/39 087 | 0.79 | 0.61, 1.03 |
| D4 | 511/100 065 | 0.89 | 0.79, 1.00 | 230/41 269 | 0.88 | 0.67, 1.14 |
| D5 | 488/110 985 | 0.76 | 0.68, 0.86 | 193/45 169 | 0.63 | 0.48, 0.83 |
| D6 | 386/88 231 | 0.77 | 0.67, 0.87 | 152/36 308 | 0.73 | 0.54, 0.99 |
| D7 | 479/103 963 | 0.81 | 0.71, 0.92 | 196/41 950 | 0.78 | 0.58, 1.04 |
| D8 | 423/102 896 | 0.74 | 0.64, 0.84 | 169/41 878 | 0.76 | 0.56, 1.05 |
| D9 | 336/96 282 | 0.63 | 0.55, 0.73 | 120/37 917 | 0.64 | 0.45, 0.91 |
| D10 | 338/100 593 | 0.62 | 0.54, 0.72 | 143/39 849 | 0.72 | 0.50, 1.03 |

#### Major depressive disorder, unspecified (F32.9)

| Deciles of fitness | Cohort analysis (N=1 013 885) | | | Sibling analysis (N=410 198) | | |
|---|---|---|---|---|---|---|
| | Cases/N | HR | 95% CI | Cases/N | HR | 95% CI |
| D1 | 3684/113 887 | Ref. | - | 1396/45 454 | Ref. | - |
| D2 | 2836/100 857 | 0.90 | 0.86, 0.94 | 1103/41 317 | 0.89 | 0.80, 1.00 |
| D3 | 2731/96 126 | 0.90 | 0.86, 0.95 | 1070/39 087 | 0.97 | 0.86, 1.08 |
| D4 | 2564/100 065 | 0.82 | 0.78, 0.86 | 1001/41 269 | 0.93 | 0.82, 1.05 |
| D5 | 2746/110 985 | 0.78 | 0.74, 0.82 | 1026/45 169 | 0.85 | 0.76, 0.96 |
| D6 | 2066/88 231 | 0.75 | 0.70, 0.79 | 813/36 308 | 0.81 | 0.71, 0.93 |
| D7 | 2330/103 963 | 0.71 | 0.67, 0.75 | 850/41 950 | 0.83 | 0.72, 0.95 |
| D8 | 2100/102 896 | 0.66 | 0.62, 0.70 | 770/41 878 | 0.71 | 0.62, 0.82 |
| D9 | 1809/96 282 | 0.61 | 0.57, 0.64 | 666/37 917 | 0.77 | 0.66, 0.89 |
| D10 | 1578/100 593 | 0.51 | 0.48, 0.55 | 613/39 849 | 0.68 | 0.58, 0.80 |

CI = confidence interval. D = decile. HR = hazard ratio. The models were adjusted for age at conscription, year of conscription, body mass index, IQ, parental education, and parental income.

**Supplemental table 8. Hazard ratios for depression diagnosis and dispensation of antidepressive medications by deciles of cardiorespiratory fitness in cohort analysis (as reported in the main article), in the sibling cohort using standard analysis, and using sibling analysis (as reported in the main article)**

| | Depression diagnosis | | |
|---|---|---|---|
| | **Cohort analysis as reported in the main article (N=1 013 885)** | **Standard analysis replicated in the sibling cohort (N=410 198)** | **Sibling analysis as reported in the main article (N=410 198)** |
| **Deciles of fitness** | **HR (95% CI)** | **HR (95% CI)** | **HR (95% CI)** |
| D1 | Ref. | Ref. | Ref. |
| D2 | 0.91 (0.88, 0.95) | 0.90 (0.85, 0.95) | 0.89 (0.82, 0.97) |
| D3 | 0.89 (0.86, 0.93) | 0.88 (0.83, 0.94) | 0.87 (0.80, 0.94) |
| D4 | 0.84 (0.81, 0.88) | 0.84 (0.79, 0.90) | 0.89 (0.81, 0.97) |
| D5 | 0.79 (0.76, 0.82) | 0.78 (0.73, 0.83) | 0.82 (0.75, 0.90) |
| D6 | 0.76 (0.73, 0.79) | 0.76 (0.71, 0.81) | 0.79 (0.71, 0.86) |
| D7 | 0.74 (0.71, 0.77) | 0.72 (0.68, 0.77) | 0.81 (0.73, 0.89) |
| D8 | 0.68 (0.65, 0.71) | 0.66 (0.62, 0.71) | 0.71 (0.64, 0.78) |
| D9 | 0.61 (0.59, 0.64) | 0.61 (0.57, 0.66) | 0.70 (0.63, 0.78) |
| D10 | 0.54 (0.52, 0.57) | 0.57 (0.53, 0.62) | 0.67 (0.59, 0.75) |
| | **Dispensation of antidepressive medications** | | |
| | **Cohort analysis as reported in the main article (N=1 013 885)** | **Standard analysis replicated in the sibling cohort (N=410 198)** | **Sibling analysis as reported in the main article (N=410 198)** |
| **Deciles of fitness** | **HR (95% CI)** | **HR (95% CI)** | **HR (95% CI)** |
| D1 | Ref. | Ref. | Ref. |
| D2 | 0.92 (0.90, 0.93) | 0.93 (0.90, 0.95) | 0.94 (0.91, 0.98) |
| D3 | 0.92 (0.90, 0.93) | 0.92 (0.90, 0.95) | 0.93 (0.90, 0.97) |
| D4 | 0.87 (0.85, 0.88) | 0.86 (0.85, 0.88) | 0.91 (0.88, 0.95) |
| D5 | 0.84 (0.82, 0.85) | 0.84 (0.84, 0.87) | 0.89 (0.86, 0.93) |
| D6 | 0.80 (0.78, 0.81) | 0.81 (0.78, 0.83) | 0.85 (0.82, 0.89) |
| D7 | 0.77 (0.76, 0.79) | 0.77 (0.75, 0.79) | 0.85 (0.81, 0.88) |
| D8 | 0.72 (0.70, 0.73) | 0.72 (0.70, 0.75) | 0.81 (0.78, 0.85) |
| D9 | 0.69 (0.68, 0.70) | 0.69 (0.67, 0.71) | 0.79 (0.76, 0.83) |
| D10 | 0.63 (0.62, 0.65) | 0.65 (0.63, 0.68) | 0.76 (0.72, 0.80) |

CI = confidence interval. D = decile. HR = hazard ratio. The models were adjusted for age at conscription, year of conscription, body mass index, IQ, parental education, and parental income.

**Supplemental table 9. Hazard ratios for depression diagnosis and dispensation of antidepressive medications by deciles of cardiorespiratory fitness in cohort and sibling analysis, restricted to those who conscribed year 1985 or later.**

| | **Depression diagnosis** | |
|---|---|---|
| | **Cohort analysis (N=439 888)** | **Sibling analysis (N=119 322)** |
| **Deciles of fitness** | **HR (95% CI)** | **HR (95% CI)** |
| D1 | Ref. | Ref. |
| D2 | 0.87 (0.83, 0.91) | 1.01 (0.78, 1.31) |
| D3 | 0.77 (0.74, 0.81) | 0.78 (0.62, 0.99) |
| D4 | 0.70 (0.67, 0.73) | 0.81 (0.64, 1.03) |
| D5 | 0.64 (0.61, 0.68) | 0.90 (0.73, 1.11) |
| D6 | 0.58 (0.55, 0.61) | 0.79 (0.64, 0.99) |
| D7 | 0.55 (0.52, 0.58) | 0.87 (0.70, 1.08) |
| D8 | 0.51 (0.48, 0.53) | 0.70 (0.56, 0.87) |
| D9 | 0.46 (0.44, 0.49) | 0.67 (0.54, 0.84) |
| D10 | 0.39 (0.37, 0.41) | 0.65 (0.52, 0.81) |
| | **Dispensation of antidepressive medications** | |
| | **Cohort analysis (N=439 888)** | **Sibling analysis (N=119 322)** |
| **Deciles of fitness** | **HR (95% CI)** | **HR (95% CI)** |
| D1 | Ref. | Ref. |
| D2 | 0.91 (0.88, 0.95) | 0.93 (0.83, 1.06) |
| D3 | 0.89 (0.85, 0.92) | 0.95 (0.85, 1.06) |
| D4 | 0.86 (0.83, 0.89) | 0.94 (0.84, 1.05) |
| D5 | 0.81 (0.78, 0.84) | 0.89 (0.81, 0.99) |
| D6 | 0.77 (0.74, 0.79) | 0.87 (0.78, 0.96) |
| D7 | 0.73 (0.71, 0.76) | 0.85 (0.77, 0.93) |
| D8 | 0.68 (0.65, 0.70) | 0.81 (0.73, 0.89) |
| D9 | 0.65 (0.63, 0.67) | 0.75 (0.68, 0.84) |
| D10 | 0.59 (0.57, 0.61) | 0.73 (0.66, 0.81) |

CI = confidence interval. D = decile. HR = hazard ratio.
The models were adjusted for age at conscription, year of conscription, body mass index, IQ, parental education, and parental income.

**Supplemental table 10. Standardised cumulative incidences of depression diagnosis and dispensation of antidepressive medications at 65 years of age by deciles of cardiorespiratory fitness in cohort and sibling analysis, allowing the effect of fitness to vary across follow-up time[a].**

### Depression diagnosis

| Deciles of fitness | Cohort analysis (N=1 013 885) | | Sibling analysis (N=410 198) | |
|---|---|---|---|---|
| | **Incidence at age 65, % (95% CI)** | **Incidence difference, % (95% CI)** | **Incidence at age 65, % (95% CI)** | **Incidence difference, % (95% CI)** |
| D1 | 8.7 (8.5, 9.0) | Ref. | 7.5 (7.0, 7.9) | Ref. |
| D2 | 8.0 (7.8, 8.3) | -0.7 (-1.0, -0.4) | 6.6 (6.2, 7.1) | -0.8 (-1.4, -0.3) |
| D3 | 7.9 (7.6, 8.1) | -0.9 (-1.2, -0.6) | 6.6 (6.2, 7.0) | -0.9 (-1.5, -0.3) |
| D4 | 7.4 (7.2, 7.6) | -1.4 (-1.7, -1.1) | 6.6 (6.2, 7.0) | -0.8 (-1.5, -0.2) |
| D5 | 6.8 (6.6, 7.2) | -1.9 (-2.2, -1.6) | 6.2 (5.8, 6.6) | -1.2 (-1.8, -0.6) |
| D6 | 6.6 (6.4, 6.9) | -2.1 (-2.4, 1.8) | 5.7 (5.3, 6.2) | -1.7 (-2.4, -1.1) |
| D7 | 6.2 (6.0, 6.4) | -2.5 (-2.9, -2.2) | 5.9 (5.4, 6.3) | -1.6 (-2.3, -0.9) |
| D8 | 5.8 (5.6, 6.0) | -3.0 (-3.3, -2.7) | 5.4 (5.0, 5.8) | -2.1 (-2.7, -1.4) |
| D9 | 5.1 (4.8, 5.3) | -3.7 (-4.0, -3.3) | 5.1 (4.6, 5.6) | -2.4 (-3.1, -1.6) |
| D10 | 4.5 (4.2, 4.7) | -4.3 (-4.6, -3.9) | 4.6 (4.1, 5.1) | -2.9 (-3.7, -2.1) |

### Dispensation of antidepressive medications

| Deciles of fitness | Cohort analysis (N=1 013 885) | | Sibling analysis (N=410 198) | |
|---|---|---|---|---|
| | **Incidence at age 65, % (95% CI)** | **Incidence difference, % (95% CI)** | **Incidence at age 65, % (95% CI)** | **Incidence difference, % (95% CI)** |
| D1 | 42.5 (42.1, 42.9) | Ref. | 37.2 (36.3, 38.1) | Ref. |
| D2 | 40.1 (39.7, 40.5) | -2.4 (-2.9, -1.9) | 35.7 (34.8, 36.5) | -1.5 (-2.6, -0.5) |
| D3 | 39.7 (39.3, 40.1) | -2.9 (-3.4, -2.3) | 35.2 (34.4, 36.1) | -2.0 (-3.1, -0.9) |
| D4 | 38.2 (37.8, 38.6) | -4.4 (4.9, -3.8) | 35.0 (34.2, 35.9) | -2.2 (-3.4, -1.0) |
| D5 | 36.8 (36.4, 37.2) | -5.7 (-6.3, -5.2) | 34.1 (33.3, 35.0) | -3.1 (-4.3, -1.9) |
| D6 | 35.6 (35.1, 36.0) | -7.0 (-7.6, -6.4) | 32.7 (31.8, 33.7) | -4.5 (-5.8, -3.2) |
| D7 | 34.1 (33.6, 34.6) | -8.5 (-9.1, -7.8) | 32.6 (31.7, 33.6) | -4.6 (-5.9, -3.2) |
| D8 | 32.3 (31.8, 32.8) | -10.3 (-10.9, -9.6) | 31.6 (30.6, 32.6) | -5.6 (-7.0, -4.2) |
| D9 | 30.6 (30.0, 31.3) | -11.9 (-12.7, -11.2) | 31.1 (29.9, 32.4) | -6.1 (-7.7, -4.4) |
| D10 | 28.9 (28.2, 29.6) | -13.6 (-14.5, -12.8) | 30.3 (28.9, 31.7) | -6.9 (-8.8, -5.1) |

[a]The incidences were allowed to vary across time using an interaction between a restricted cubic spline with three degrees of freedom of the follow-up time (centile 33 and 67 of the distribution of the uncensored log survival times) and the fitness deciles.

CI = confidence interval. D = decile. HR = hazard ratio. The models were adjusted for age at conscription, year of conscription, body mass index, IQ, parental education, and parental income, and cardiorespiratory fitness was modelled as a time-dependent exposure.

**Supplemental table 11. Hazard ratios for depression diagnosis and dispensation of antidepressive medications by deciles of cardiorespiratory fitness expressed as Wmax (as reported in the main article), Wmax/kg, or estimated $VO_2$max in cohort and sibling analysis.**

**Depression diagnosis**

| | **Wmax (as reported in main article)** | | **Wmax/kg** | | **Estimated $VO_2$max** | |
|---|---|---|---|---|---|---|
| | **Cohort analysis (N=1 013 885)** | **Sibling analysis (N=410 198)** | **Cohort analysis (N=1 013 885)** | **Sibling analysis (N=410 198)** | **Cohort analysis (N=1 013 885)** | **Sibling analysis (N=410 198)** |
| **Deciles of fitness** | **HR (95% CI)** | **HR (95% CI)** | **HR (95% CI)** | **HR (95% CI)** | **HR (95% CI)** | **HR (95% CI)** |
| D1 | Ref. | Ref. | Ref. | Ref. | Ref. | Ref. |
| D2 | 0.91 (0.88, 0.95) | 0.89 (0.82, 0.97) | 0.90 (0.86, 0.93) | 0.87 (0.80, 0.95) | 0.90 (0.86, 0.93) | 0.87 (0.80, 0.95) |
| D3 | 0.89 (0.86, 0.93) | 0.87 (0.80, 0.94) | 0.85 (0.82, 0.89) | 0.89 (0.81, 0.97) | 0.85 (0.82, 0.89) | 0.89 (0.81, 0.97) |
| D4 | 0.84 (0.81, 0.88) | 0.89 (0.81, 0.97) | 0.83 (0.79, 0.86) | 0.83 (0.75, 0.91) | 0.83 (0.79, 0.86) | 0.83 (0.75, 0.91) |
| D5 | 0.79 (0.76, 0.82) | 0.82 (0.75, 0.90) | 0.80 (0.77, 0.83) | 0.82 (0.75, 0.90) | 0.80 (0.77, 0.83) | 0.82 (0.75, 0.90) |
| D6 | 0.76 (0.73, 0.79) | 0.79 (0.71, 0.86) | 0.76 (0.73, 0.79) | 0.79 (0.72, 0.87) | 0.76 (0.73, 0.79) | 0.79 (0.72, 0.87) |
| D7 | 0.74 (0.71, 0.77) | 0.81 (0.73, 0.89) | 0.73 (0.70, 0.76) | 0.73 (0.66, 0.81) | 0.73 (0.70, 0.76) | 0.73 (0.66, 0.81) |
| D8 | 0.68 (0.65, 0.71) | 0.71 (0.64, 0.78) | 0.69 (0.66, 0.72) | 0.76 (0.69, 0.85) | 0.69 (0.66, 0.72) | 0.76 (0.69, 0.85) |
| D9 | 0.61 (0.59, 0.64) | 0.70 (0.63, 0.78) | 0.65 (0.62, 0.68) | 0.70 (0.63, 0.78) | 0.65 (0.62, 0.68) | 0.70 (0.63, 0.78) |
| D10 | 0.54 (0.52, 0.57) | 0.67 (0.59, 0.75) | 0.57 (0.54, 0.60) | 0.69 (0.61, 0.78) | 0.57 (0.54, 0.60) | 0.69 (0.61, 0.78) |
| **Cardiorespiratory fitness, per 1 MET** | | | | | 0.93 (0.93, 0.94) | 0.95 (0.94, 0.96) |

**Dispensation of antidepressive medications**

| | **Wmax (as reported in main article)** | | **Wmax/kg** | | **Estimated $VO_2$max** | |
|---|---|---|---|---|---|---|
| | **Cohort analysis (N=1 013 885)** | **Sibling analysis (N=410 198)** | **Cohort analysis (N=1 013 885)** | **Sibling analysis (N=410 198)** | **Cohort analysis (N=1 013 885)** | **Sibling analysis (N=410 198)** |
| **Deciles of fitness** | **HR (95% CI)** | **HR (95% CI)** | **HR (95% CI)** | **HR (95% CI)** | **HR (95% CI)** | **HR (95% CI)** |
| D1 | Ref. | Ref. | Ref. | Ref. | Ref. | Ref. |
| D2 | 0.92 (0.90, 0.93) | 0.94 (0.91, 0.98) | 0.91 (0.90, 0.93) | 0.95 (0.92, 0.99) | 0.91 (0.90, 0.93) | 0.95(0.92, 0.99) |
| D3 | 0.92 (0.90, 0.93) | 0.93 (0.90, 0.97) | 0.88 (0.87, 0.90) | 0.92 (0.89, 0.96) | 0.88 (0.87, 0.90) | 0.92 (0.89, 0.96) |
| D4 | 0.87 (0.85, 0.88) | 0.91 (0.88, 0.95) | 0.84 (0.83, 0.86) | 0.89 (0.86, 0.93) | 0.84 (0.83, 0.86) | 0.89 (0.86, 0.93) |
| D5 | 0.84 (0.82, 0.85) | 0.89 (0.86, 0.93) | 0.81 (0.79, 0.82) | 0.86 (0.82, 0.90) | 0.81 (0.79, 0.82) | 0.86 (0.82, 0.90) |
| D6 | 0.80 (0.78, 0.81) | 0.85 (0.82, 0.89) | 0.80 (0.79, 0.82) | 0.86 (0.82, 0.90) | 0.80 (0.79, 0.82) | 0.86 (0.82, 0.90) |
| D7 | 0.77 (0.76, 0.79) | 0.85 (0.81, 0.88) | 0.76 (0.74, 0.77) | 0.85 (0.81, 0.89) | 0.76 (0.74, 0.77) | 0.85 (0.81, 0.89) |
| D8 | 0.72 (0.70, 0.73) | 0.81 (0.78, 0.85) | 0.73 (0.71, 0.74) | 0.82 (0.79, 0.86) | 0.73 (0.71, 0.74) | 0.82 (0.79, 0.86) |
| D9 | 0.69 (0.68, 0.70) | 0.79 (0.76, 0.83) | 0.69 (0.68, 0.70) | 0.79 (0.75, 0.83) | 0.69 (0.68, 0.70) | 0.79 (0.75, 0.83) |
| D10 | 0.63 (0.62, 0.65) | 0.76 (0.72, 0.80) | 0.64 (0.63, 0.65) | 0.78 (0.74, 0.82) | 0.64 (0.63, 0.65) | 0.78 (0.74, 0.82) |
| **Cardiorespiratory fitness, per 1 MET** | | | | | 0.94 (0.94, 0.94) | 0.96 (0.96, 0.97) |

CI = confidence interval. D = decile. HR = hazard ratio. MET = metabolic equivalent of task, computed from dividing estimated $VO_2$max/ 3.5.
The models were adjusted for age at conscription, year of conscription, body mass index, IQ, parental education, and parental income.
